# DeepDrug: An Expert-led Domain-specific AI-Driven Drug-Repurposing Mechanism for Selecting the Lead Combination of Drugs for Alzheimer’s Disease

**DOI:** 10.1101/2024.07.06.24309990

**Authors:** Victor OK Li, Yang Han, Tushar Kaistha, Qi Zhang, Jocelyn Downey, Illana Gozes, Jacqueline CK Lam

**Affiliations:** Department of Electrical and Electronic Engineering, The University of Hong Kong, Hong Kong; Department of Human Molecular Genetics and Biochemistry, Sackler Faculty of Medicine, Tel Aviv University, Israel

**Keywords:** DeepDrug, Alzheimer’s Disease, expert-led AI drug-repurposing, graph neural network, somatic and germline mutations, long genes, lead combination of AD drugs, directed biomedical graph, neuro-degenerative, inflammation, immunological, aging pathways, and pathway convergence

## Abstract

Alzheimer’s Disease (AD) significantly aggravates human dignity and quality of life. While newly approved amyloid immunotherapy has been reported, effective AD drugs remain to be identified. Here, we propose a novel AI-driven drug-repurposing method, DeepDrug, to identify a lead combination of approved drugs to treat AD patients. DeepDrug advances drug-repurposing methodology in four aspects. Firstly, it incorporates expert knowledge to extend candidate targets to include long genes, immunological and aging pathways, and somatic mutation markers that are associated with AD. Secondly, it incorporates a signed directed heterogeneous biomedical graph encompassing a rich set of nodes and edges, and node/edge weighting to capture crucial pathways associated with AD. Thirdly, it encodes the weighted biomedical graph through a Graph Neural Network into a new embedding space to capture the granular relationships across different nodes. Fourthly, it systematically selects the high-order drug combinations via diminishing return-based thresholds. A five-drug lead combination, consisting of Tofacitinib, Niraparib, Baricitinib, Empagliflozin, and Doxercalciferol, has been selected from the top drug candidates based on DeepDrug scores to achieve the maximum synergistic effect. These five drugs target neuroinflammation, mitochondrial dysfunction, and glucose metabolism, which are all related to AD pathology. DeepDrug offers a novel AI-and-big-data, expert-guided mechanism for new drug combination discovery and drug-repurposing across AD and other neuro-degenerative diseases, with immediate clinical applications.

## Introduction

Alzheimer’s disease (AD) significantly deteriorates human health and quality of life^1–8^. Globally, around 50 million people have suffered from AD and related forms of dementia, generating 28.8 million disability-adjusted life-years^1^. In general, drug development is a hugely expensive endeavor. It takes USD$2.6 billion for a candidate drug to go through the developmental process prior to regulatory approval. At present, no effective disease-modifying treatment or preventative therapies have been found, while the process of screening effective AD drug candidates has remained lengthy and data-constrained^9–13^. Only six drugs and one drug combination comprising two of the six drugs have been approved by the US Food and Drug Administration (FDA) for AD treatment. Among these, only one has been reported to postpone cognitive decline, with the rest targeting at the symptoms rather than the root causes of AD.

In our study, instead of focusing individually on cholinergic deficit, excitotoxicity, or amyloid deposition, the key hallmarks of AD encompassing specific pharmaceutical intervention, an artificial intelligence (AI)-based holistic approach for multi-drug repurposing has been targetted. Drug-repurposing is a strategy that identifies new uses for approved drugs. It sheds new hopes for the discovery of treatments for AD and other neurodegenerative diseases, by repurposing existing clinical or non-clinical drugs reported in the drug banks^14–21^. As such, network-based statistical methods were proposed to identify putative therapeutics that can be repurposed for cancer, based on the distance (the shortest path in the network) between the drug targets (proteins) and disease proteins of cancer, assuming that the mutated genes that generate the disease proteins are connected with cancer through protein-protein interactions^22^. Moreover, drug combinations, which consist of more than one drug with different mechanisms of action and in a single dosage form, were reported to increase the chance of effective therapy^23,24^. Statistical methods were previously used to identify repurposed two-drug combinations for hypertension and cancer by exploring the network-based relationship between two drugs (and their targets) and disease proteins^25^. Given the large search space of high-order drug combinations, a random forest was proposed to identify effective drug combinations (from three to five drugs) for tuberculosis therapy based on pairwise drug response data only^26^. More recently, network-based drug repurposing has been exploited to combat COVID-19 infection. Belyaeva et al. (2021)^27^ integrated multi-omics data with aging signatures into a causal network model to identify drug candidates to treat COVID-19. Howell et al. (2022)^28^ constructed a virus-host interactome as a discrete qualitative network and used node perturbations to identify two-drug combinations for different stages of COVID-19 infection. Further, Gysi et al. (2021) developed a graph neural network (GNN) for COVID-19 treatment recommendations based on a link prediction problem on a biomedical graph consisting of nodes (drugs, proteins, diseases) and edges (protein-protein interactions, drug-target associations, disease-protein associations, and drug-disease indications)^29^. Hsieh et al. (2021)^30^ developed a GNN model to encode graph nodes (drugs and proteins) into a lower-dimensional space and determined COVID-19 treatment based on the closeness of the drugs to COVID-19 disease proteins^30^.

In the context of AD drug-repurposing, a few studies have demonstrated several successful applications of computational drug-repurposing methods. Siavelis et al. (2016)^31^ combined a network-based approach with traditional statistical measures, capitalizing on the results obtained from an AD-related microarray study to demonstrate that genes are differentially expressed when AD-related cells are exposed to a test panel of drugs. Taubes et al. (2021)^32^ performed computational AD drug-repurposing based on transcriptomic perturbations of drugs best reversing APOE4-related AD signatures, in combination with experimental evidence observed in mouse models, to demonstrate the efficacy of bumetanide in modulating pathways associated with APOE4, a major genetic risk factor for AD. Bayraktar et al. (2023)^33^ identified glutaminase as an AD drug target and performed computational drug-repurposing to reveal drugs that can reduce glutamate production based on gene expression data. Furthermore, multi-omics data and systems biology approaches have become increasingly adopted in AD drug-repurposing. Zhou et al. (2021)^34^ integrated genetic, transcriptomic, and proteomic data to identify potential drug targets for AD treatment through network-based statistical analysis. Azizan et al. (2024)^35^ explored the molecular links between metabolic disorders and AD through a systems biology approach, demonstrating the common pathways and molecular targets shared by metabolic conditions (hypertension, non-alcoholic fatty liver disease, and diabetes) and AD, and providing new insights into AD pathophysiology and drug-repurposing. Even though these computational approaches are yet to exploit the full potential of AI in capturing the complex relationships of high-dimensional biomedical data, they have demonstrated the significance of synthesizing diverse biological data sources to uncover the underlying disease mechanisms and improve the efficacy of drug-repurposing for AD.

More recently, the landscape of AD drug-repurposing has rapidly evolved given the advancement in AI-driven computational methods^36,37^. Rodriguez et al. (2021) developed a machine learning framework to predict a list of genes that associate with different stages of AD, based on gene expression data from multiple datasets, for drug repurposing. Ghiam et al. (2022)^38^ examined the role of long non-coding RNAs (lncRNAs) in the cross-talk between diabetes and AD using two bipartite networks, including (1) an mRNA-miRNA network with common mRNAs in diabetes and AD and (2) an miRNA-lncRNA network with high-degree miRNAs obtained from the mRNA-miRNA network. They utilized machine learning to predict the effects of drugs on the expression of high-degree lncRNAs selected from the miRNA-lncRNA network. Furthermore, a few studies have utilized GNNs to model the complex interactions between different entities within biological networks, integrating various biomedical datasets to address the complexity and heterogeneity of AD. Zeng et al. (2019)^39^ proposed DeepDR, a GNN model to learn high-level representations of drugs and diseases from biomedical networks and predict drug-disease associations. Xu et al. (2022)^40^ proposed NETTAG, integrating genome-wide association studies (GWAS) findings and multi-omics data to predict AD-risk genes and identify druggable targets, with the help of a modified GNN model capturing the sparse structure and clustering of a protein-protein interaction (PPI) network. Pan et al. (2023)^41^ developed AI-DrugNet to learn the representation of individual drug-target pair, using a GNN model based on a drug-target pair network and predict drug combinations for AD based on the combined features of two drug-target pairs. Hsieh et al. (2023)^42^ further used a graph autoencoder model to learn node representations from an AD knowledge graph consisting of four node types (drugs, genes, pathways, and gene ontology) and their interactions, identifying top-ranking drugs according to multi-level drug efficacy evidence, ranging from transcriptomic patterns to clinical trials, and two-drug combinations selected from top-ranking drugs with synergistic effects in targeting multiple disease pathways.

Here, we propose DeepDrug, an expert-led AI-driven drug-repurposing framework, to identify a lead combination of approved drugs for AD treatment. While the underlying mechanism of AD remains largely unknown^12,43–45^, the novel DeepDrug model features the incorporation of domain-specific knowledge from neuroscience experts into AI-driven drug-repurposing. Specifically, DeepDrug has four novelties as compared to previous studies: (i) It integrates expert-led knowledge (including long genes, immunological and aging pathways, and somatic mutation markers identified from the blood and the brain with AD) to select candidate genes and drugs relevant to AD, using their GNN-based high-level representations to calculate drug-gene scores. In contrast, previous GNN-based approaches focused primarily on drug-disease association^39^, protein-protein interaction^40^, or drug-target interaction^41^; (ii) It generates an expert-led signed directed heterogeneous biomedical graph to encompass a rich set of nodes and edges, carefully incorporating expert knowledge in graph construction to capture crucial pathways that associate with AD. The expert-led biomedical graph distinguishes itself from previous AD biomedical graphs^42^, allowing (a) nodes/edges weight assignments and (b) positive/negative directed edges, better capturing AD domain-specific knowledge; (iii) It encodes the expert-led biomedical graph through a signed directed GNN into a new embedding space to better capture the rich relationships between different types of nodes, deviating from traditional statistical measures relying solely on the shortest path metric based on signed directed biomedical graphs^46^ or recent graph representation learning models^42^ that do not account for the weights of nodes/edges and the signs/directions of edges. Building on top of the domain-specific node/edge weight assignments, such as drug-target affinity, and the signed and directed edges, such as drug-target activation or inhibition, DeepDrug can further enhance the accuracy of prediction of successful drug candidates; (iv) It optimizes the selection of repurposed drug-combinations systematically, prioritizing single drug candidates via diminishing return-determined thresholds and identifying the lead drug combinations beyond two drugs^41,42^.

Our DeepDrug drug-repurposing framework consists of a three-stage methodology. Firstly, the construction of a heterogeneous directed biomedical graph consisting of interconnected genes, proteins, and drugs to capture the network characteristics of AD pathology, accounting for the known associations across the expert-led domain-specific AD pathways. Secondly, the biomedical graph is taken as the input to an artificial intelligence (AI)-driven GNN framework, which creates the embeddings of the drug and gene nodes in a lower dimensional embedding space as the outputs. Thirdly, a drug scoring and selection analysis is conducted to generate the drug-gene scores and identify a lead combination of repurposed AD drug candidates with the top drug-gene scores for clinical verification. A five-drug combination, consisting of Tofacitinib, Niraparib, Diltiazem, Pantoprazole, and Pravastatin, has been selected from the top repurposed drugs to achieve the maximum synergistic effect. These five drugs target neuroinflammation, mitochondrial dysfunction, and cholesterol metabolism, which are all related to AD pathology. Figure 1 summarizes our proposed DeepDrug methodology and novelties.

**Figure 1.**
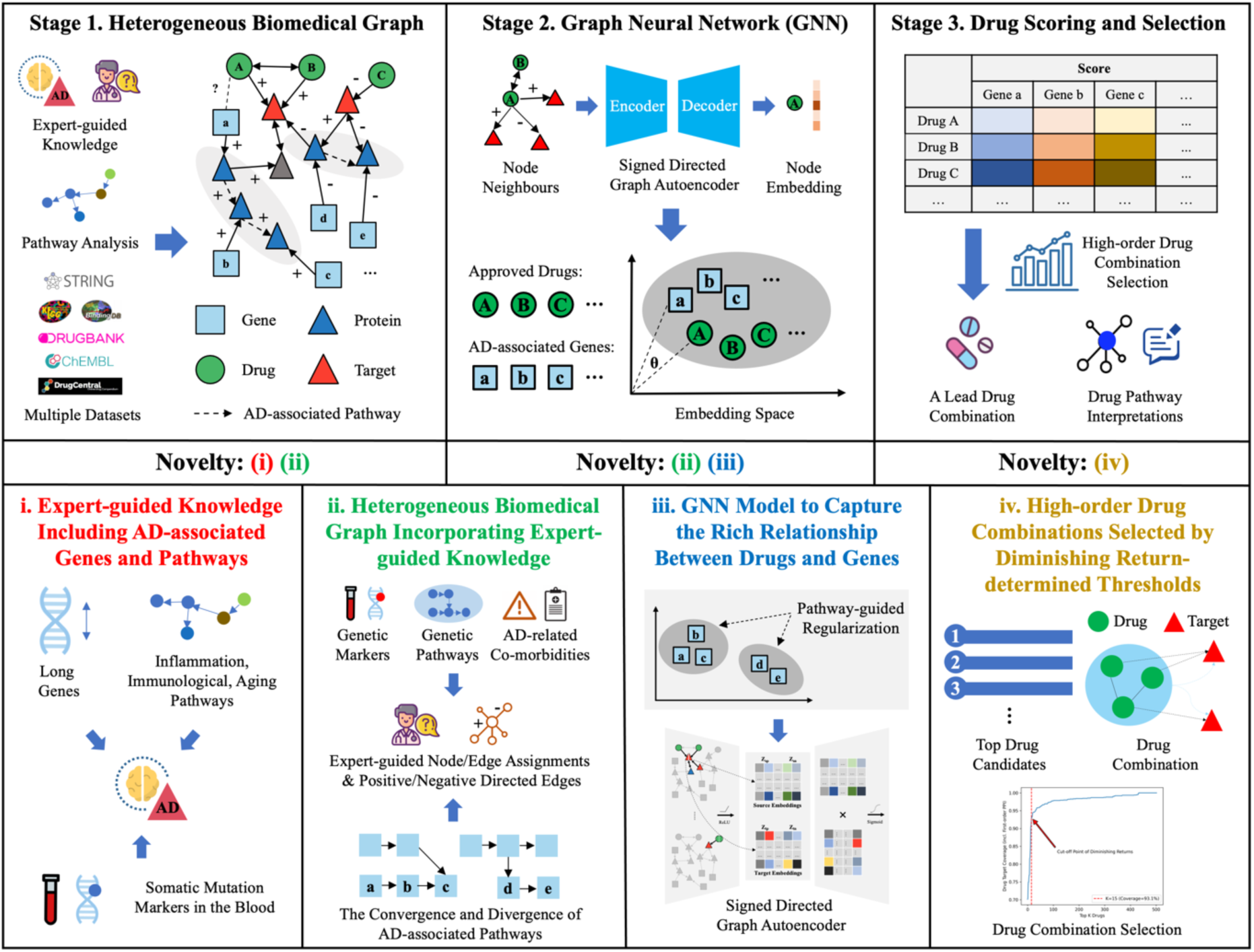
DeepDrug Framework and Novelties

## Results

### Identification of the Top Approved Drug Candidates from DeepDrug

By means of DeepDrug methodology (see “Methods”), we have generated a list of top repurposed approved drugs as well as a lead combination of five repurposed drugs for AD treatment. Table 1 shows the top 15 drug candidates and their respective scores based on the findings generated by DeepDrug (see “Scoring and Selection of Drug and Drug Combination”). All these drugs are potentially relevant candidates for AD treatment based on evidence from previous literature. Specifically, the top drug, Tofacitinib, was identified as one of the top repurposed drugs in AI-driven AD drug-repurposing research,^47^ subsequently tested in mouse models^48^. Among the top five drugs, three were Janus kinase (JAK) inhibitors, which can potentially reduce neuroinflammation, an important factor in AD pathology^49^. The other two anticancer drugs, Niraparib and Palbociclib, were considered potential agents for AD drug-repurposing targeting autophagy^50^. Among the remaining ten drugs, Sirolimus, a mammalian target of rapamycin (mTOR) inhibitor, was considered a potential therapeutic agent given its role in autophagy^51^. Empagliflozin, Febuxostat, Doxercalciferol, and Zinc Acetate can be relevant candidates for AD treatment because they were formerly approved for treating AD-associated comorbidities or risk factors, including diabetes^52^, gout^53^, vitamin D deficiency^54^, and zinc deficiency^55^. Raloxifene was previously identified to treat women with AD^56^, a subpopulation that carries a higher risk of AD. Valproic Acid and Olanzapine were formerly approved for treating neurological diseases such as seizures and schizophrenia and were identified as promising agents for AD treatment^57,58^. Other candidates, including Adenosine^59^ and Miconazole^60^, carry the potentials to reduce neuroinflammation responses.

**Table 1.** Top Repurposed Drug Candidates for AD by DeepDrug.

| DeepDrug Ranking | DeepDrug Score | Drug | Approved Treatment | Description |
| --- | --- | --- | --- | --- |
| 1 | 0.2692 | Tofacitinib | Rheumatic conditions | Janus kinase (JAK) inhibitor |
| 2 | 0.1528 | Niraparib | Cancer | Poly-ADP ribose polymerase (PARP) inhibitor |
| 3 | 0.1454 | Palbociclib | Cancer | Cyclin-dependent kinase 4/6 (CDK4/6) inhibitor |
| 4 | 0.1446 | Filgotinib | Rheumatic conditions | JAK inhibitor |
| 5 | 0.1436 | Baricitinib | Rheumatic conditions | JAK inhibitor |
| 6 | 0.1318 | Sirolimus | Organ transplant rejection | Mammalian target of rapamycin (mTOR) inhibitor |
| 7 | 0.1195 | Empagliflozin | Diabetes | Sodium-glucose co-transporter-2 (SGLT2) inhibitor |
| 8 | 0.1096 | Febuxostat | Hyperuricemia<br>Gout | Xanthine oxidase (XO) inhibitor |
| 9 | 0.1084 | Raloxifene | Osteoporosis | Estrogen receptor modulator (SERM) |
| 10 | 0.0964 | Doxercalciferol | Hyperparathyroidism | Vitamin D2 analog |
| 11 | 0.0939 | Valproic Acid | Seizures | Fatty acid derivative and anticonvulsant |
| 12 | 0.0938 | Miconazole | Fungal infections | Azole antifungal |
| 13 | 0.0937 | Zinc Acetate | Zinc deficiency<br>Wilson's disease | Zinc |
| 14 | 0.0936 | Adenosine | Supraventricular tachycardia | Antiarrhythmic agent |
| 15 | 0.0917 | Olanzapine | Schizophrenia<br>Bipolar I disorder | Antipsychotic agent |

### Identification of a Lead Combination of Drugs Based on the Top Approved Drugs

Based on the 15 drug candidates identified, drug combinations, ranging from two-drug to six-drug combinations, have been generated by DeepDrug (see “Drug Combination Optimisation”). Figure 2 shows the distribution of the drug combination scores from two-drug combinations up to six-drug combinations.

**Figure 2.**
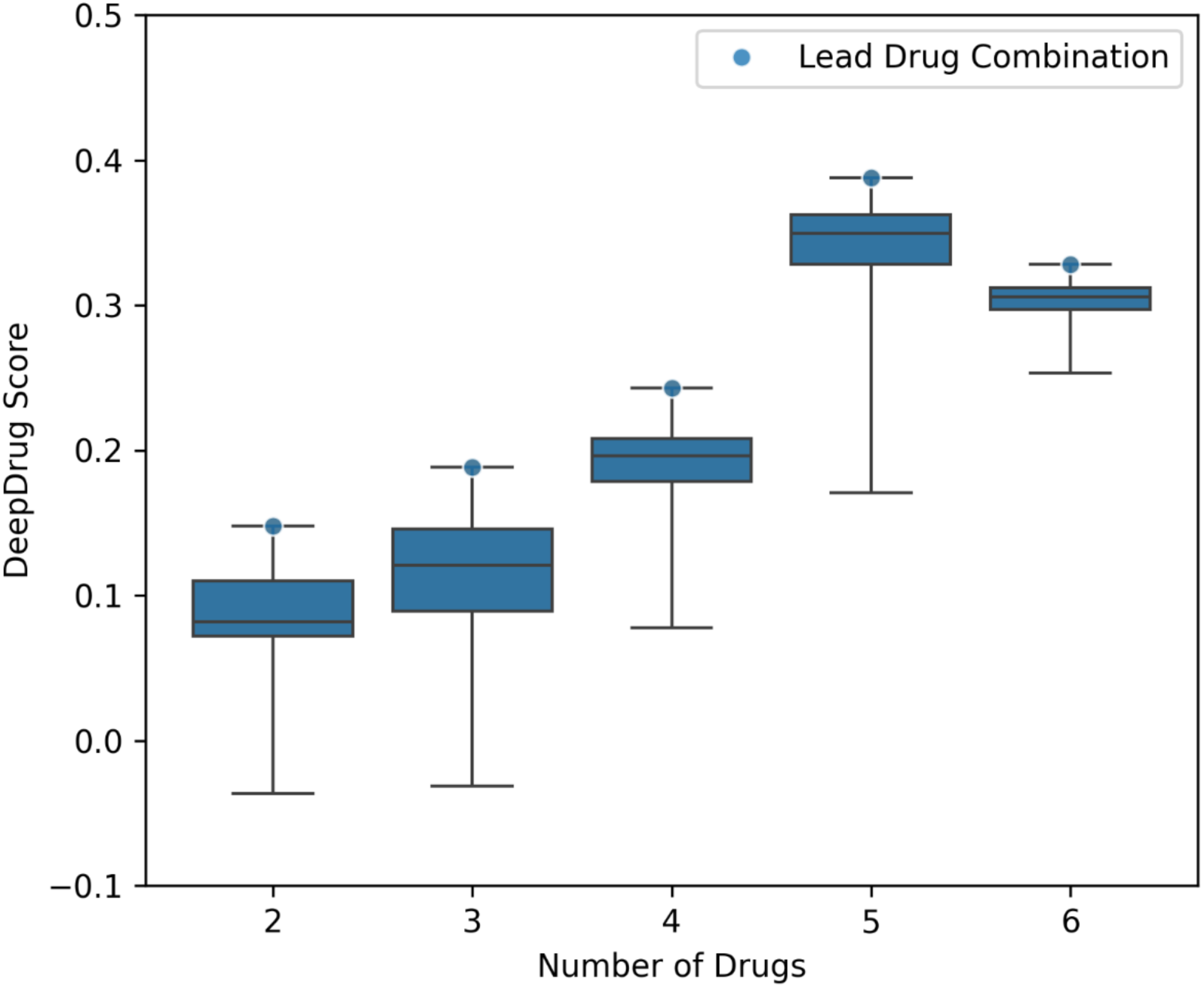
Box Plot of Drug Combination Scores [Note: This figure shows the scores of all drug combinations, from two-drug combinations up to six-drug combinations, grouped by the number of drugs in the drug combination, including the minimum, 25^th^ percentile, median, 75^th^ percentile, and maximum scores. The leading drug combination has been selected from the top five-drug combinations, which is the optimal number of drugs in drug combinations based on the cut-off point where the maximum score has been reached.]

The top ten five-drug combinations for AD among 3,003 combinations (choosing 5 from 15 drugs) are shown in Table 2. The small variations in the top five-drug combination scores align with previous experimental and simulation results, which suggest that an effective high-order drug combination is not “a needle in a haystack” and that many combinations of novel agents may enhance efficacy^61^. Moreover, although Valproic Acid and Miconazole have not been found in the top ten drugs (though they are in the top 15), they can be found in the top ten five-drug combination list, potentially due to their complementary interactions with other drugs. For example, Valproic Acid can reduce the expression of pro-inflammatory cytokines through its histone deacetylase (HDAC) inhibition^62^, while the other top drugs, including JAK inhibitors such as Tofacitinib and Baricitinib, can further regulate cytokine signaling^49^, potentially providing a synergistic reduction in neuroinflammation and a more comprehensive treatment strategy for AD.

**Table 2.** The Top Ten Five-drug Combinations for AD.

| Drug 1 | Drug 2 | Drug 3 | Drug 4 | Drug 5 | DeepDrug Score |
| --- | --- | --- | --- | --- | --- |
| Tofacitinib | Niraparib | Baricitinib | Empagliflozin | Doxercalciferol | 0.3876 |
| Tofacitinib | Niraparib | Empagliflozin | Doxercalciferol | Valproic acid | 0.3857 |
| Tofacitinib | Palbociclib | Empagliflozin | Doxercalciferol | Valproic acid | 0.3847 |
| Tofacitinib | Niraparib | Sirolimus | Doxercalciferol | Miconazole | 0.3844 |
| Tofacitinib | Niraparib | Palbociclib | Empagliflozin | Doxercalciferol | 0.3843 |
| Tofacitinib | Baricitinib | Empagliflozin | Doxercalciferol | Valproic acid | 0.3842 |
| Tofacitinib | Palbociclib | Empagliflozin | Febuxostat | Valproic acid | 0.3839 |
| Tofacitinib | Palbociclib | Sirolimus | Valproic acid | Miconazole | 0.3837 |
| Tofacitinib | Niraparib | Baricitinib | Sirolimus | Miconazole | 0.3836 |
| Tofacitinib | Baricitinib | Empagliflozin | Febuxostat | Valproic acid | 0.3836 |

The lead five-drug combination selected using DeepDrug comprises Tofacitinib, Niraparib, Baricitinib, Empagliflozin, and Doxercalciferol. The mechanism of these five drugs based on publicly available information on DrugBank and how these drugs are involved in different stages of AD drug discovery are listed in Table 3. These drugs were previously approved to treat rheumatic conditions, cancer, diabetes, and hyperparathyroidism. As shown in Table 3, each of the five drug candidates in the lead drug combination is relevant for inhibiting the AD pathology. Specifically, Tofacitinib is a JAK inhibitor targeting the JAK-STAT pathway, which promotes neuroinflammation responses mediated by immune cell types such as microglia and astrocytes^49^. Microglia dysfunction characterized by proinflammatory cytokines is related to the clearance/accumulation of amyloid beta, contributing to AD pathology^63^. Niraparib inhibits PARP enzymes, which may contribute to neurodegeneration through metabolic impairment and cell death regulation^64^. Baricitinib is also a JAK inhibitor, which can reinforce Tofacitinib by inhibiting JAK and reducing the production of these cytokines, potentially attenuating neuroinflammatory processes. Empagliflozin is an SGLT2 inhibitor targeting the shared pathways between diabetes and AD, including the insulin signaling pathway^65^. Doxercalciferol is a Vitamin D analog triggering neuronal protection actions against inflammatory and oxidative stress^66^. Moreover, the relevance of these five drug candidates in AD pathology has been demonstrated by their individual involvements in different stages of AD drug discovery; whilst a cited review highlighted the use of Niraparib for AD drug-repurposing^50^, an epidemiological study linked Doxercalciferol to reduced AD risk, based on observational medical records data^67^ and animal models that used Tofacitinib and Empagliflozin to treat AD in mice^48,65^, an interventional human clinical trial (Phase II) investigated the effectiveness of Baricitinib on AD treatment^68^.

**Table 3.** A Lead Five-drug Combination of FDA-Approved Drugs for AD.

| **Drug** | **Tofacitinib** | **Niraparib** | **Baricitinib** | **Empagliflozin** | **Doxercalciferol** |
| --- | --- | --- | --- | --- | --- |
| **Description** | JAK inhibitor | PRAP inhibitor | JAK inhibitor | SGLT2 inhibitor | Vitamin D2 analog |
| **Target Disease** | Rheumatic conditions | Cancer | Rheumatic conditions | Diabetes | Hyperparathyroidism |
| **AD Drug Discovery** | Mouse model<sup>48</sup> | Literature review<sup>50</sup> | Clinical trial (Phase II)<sup>68</sup> | Mouse model<sup>65</sup> | Epidemiological study<sup>67</sup> |
| **Mechanism** | Tofacitinib is a JAK inhibitor that prevents the body from responding to cytokine signals, inhibiting the JAK-STAT pathway to decrease the inflammatory response. | Niraparib is a potent and selective inhibitor of PARP enzymes, which plays an important role in DNA repair. | Baricitinib inhibits JAK proteins, modulates interleukin, interferon, and growth factor signaling pathways, reduces JAK1/JAK2 expression in mutated cells, and induces cell apoptosis. | Empagliflozin is a potent inhibitor of renal SGLT2 transporters located in the proximal tubules of the kidneys and works to lower blood glucose levels via an increase in glucosuria. | Doxercalciferol is a synthetic Vitamin D2 analog; its biologically active form controls the absorption and mobilization of calcium in the body. |

Figure 3 visualizes the lead five-drug combination in the embedding space of the signed directed biomedical graph to understand how the lead combination can work together for AD treatment. The projection is based on the t-SNE technique, which maps high-dimensional data into a three-dimensional scatter plot^69^. The AD-associated gene embeddings and the reversed drug embeddings are shown in Figure 3, where a shorter distance between a drug and a gene in the embedding space suggests a higher chance that the drug can “treat” the gene, either by inhibiting the AD-risk protein or by promoting the AD-protective protein. On average, the five drugs in the lead combination are closer to the AD-associated genes. These five drugs can be grouped into three clusters, and the three clusters are distributed across different parts of the embedding space. This pattern suggests that some drugs can reinforce others while others can be complementary. Specifically, Tofacitinib (JAK inhibitor) and Baricitinib (JAK inhibitor) are the two closest drugs in the embedding space when compared to other drugs, suggesting a strong reinforcement effect that addresses dysfunction within immunological pathways, given that both drugs can target neuroinflammation-related pathways (in particular, the JAK-STAT pathway^49^). Moreover, Niraparib (PARP inhibitor) and Doxercalciferol (vitamin D2 analog) are closer in the embedding space. One possible explanation is that the active form of vitamin D may also inhibit PARP^70^, and the two drugs may provide anti-inflammatory action and reduced oxidative stress through PARP inhibition and vitamin D pathways^66,71^. In contrast, Empagliflozin (SGLT2 inhibitor) is distant from the remaining four drugs. One possible explanation being that Empagliflozin may play an independent complementary role in AD treatment, by targeting the insulin signaling pathway related to glucose, lipid, and energy homeostasis^65^.

**Figure 3.**
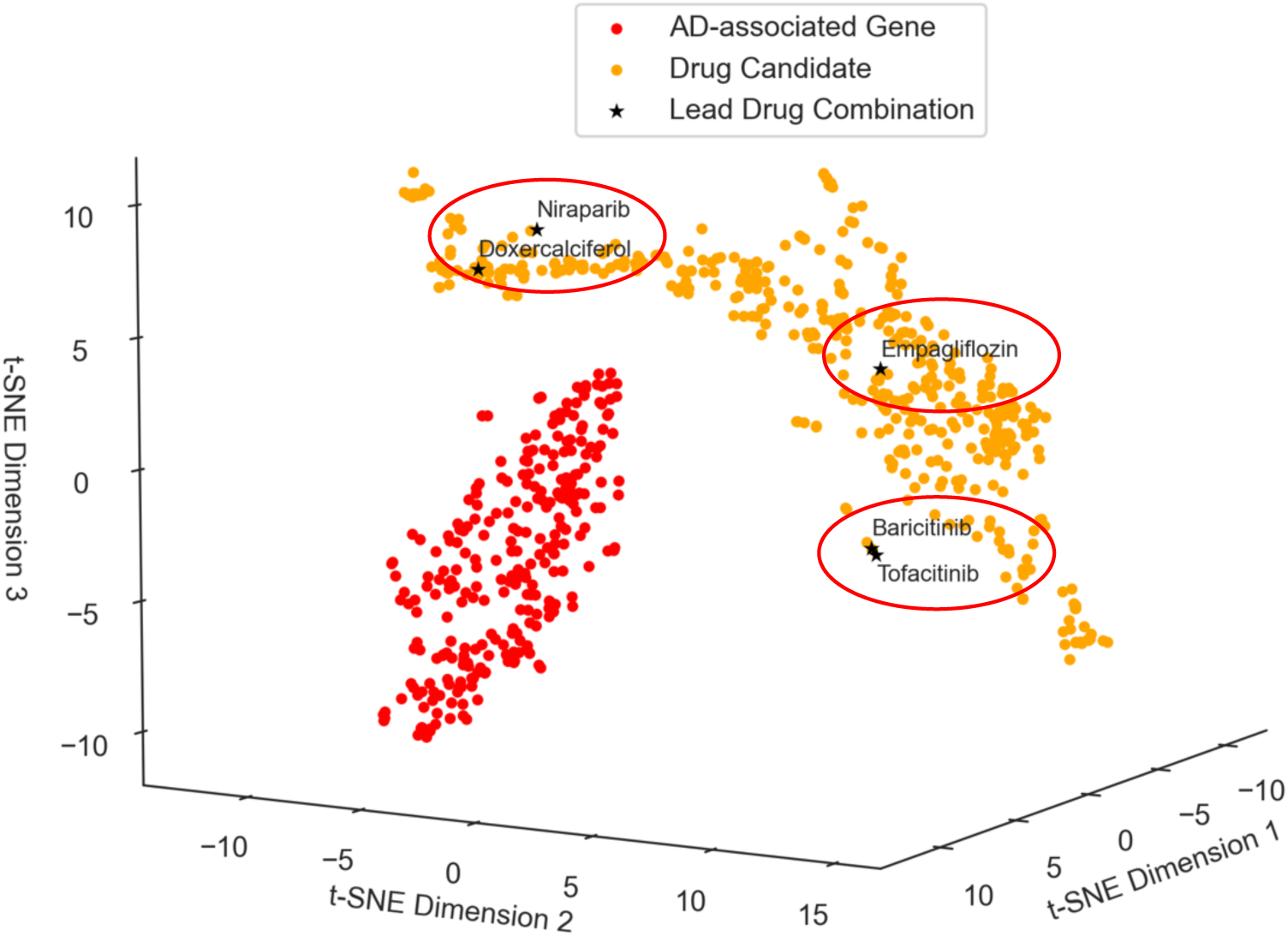
DeepDrug Prediction of the Five-drug Combination in the Embedding Space

Figure 4 further illustrates the potential mechanism and biological relevance of the lead five-drug combination in AD treatment using network and pathway analysis. Specifically, Figure 4 shows the protein targets of the lead combination (which achieves the highest DeepDrug score), and how these targets are linked to the top ten AD-associated genes (among the 310 genes) via the shortest paths of individual biomedical graphs of a drug-target-gene interaction network. Although the top genes involved in the KEGG AD pathway (such as APP, APOE, PSEN1, and MAPT) are not directly targeted by the five drugs in combination, these drugs can still target the AD-associated genes through the PPI network.

**Figure 4.**
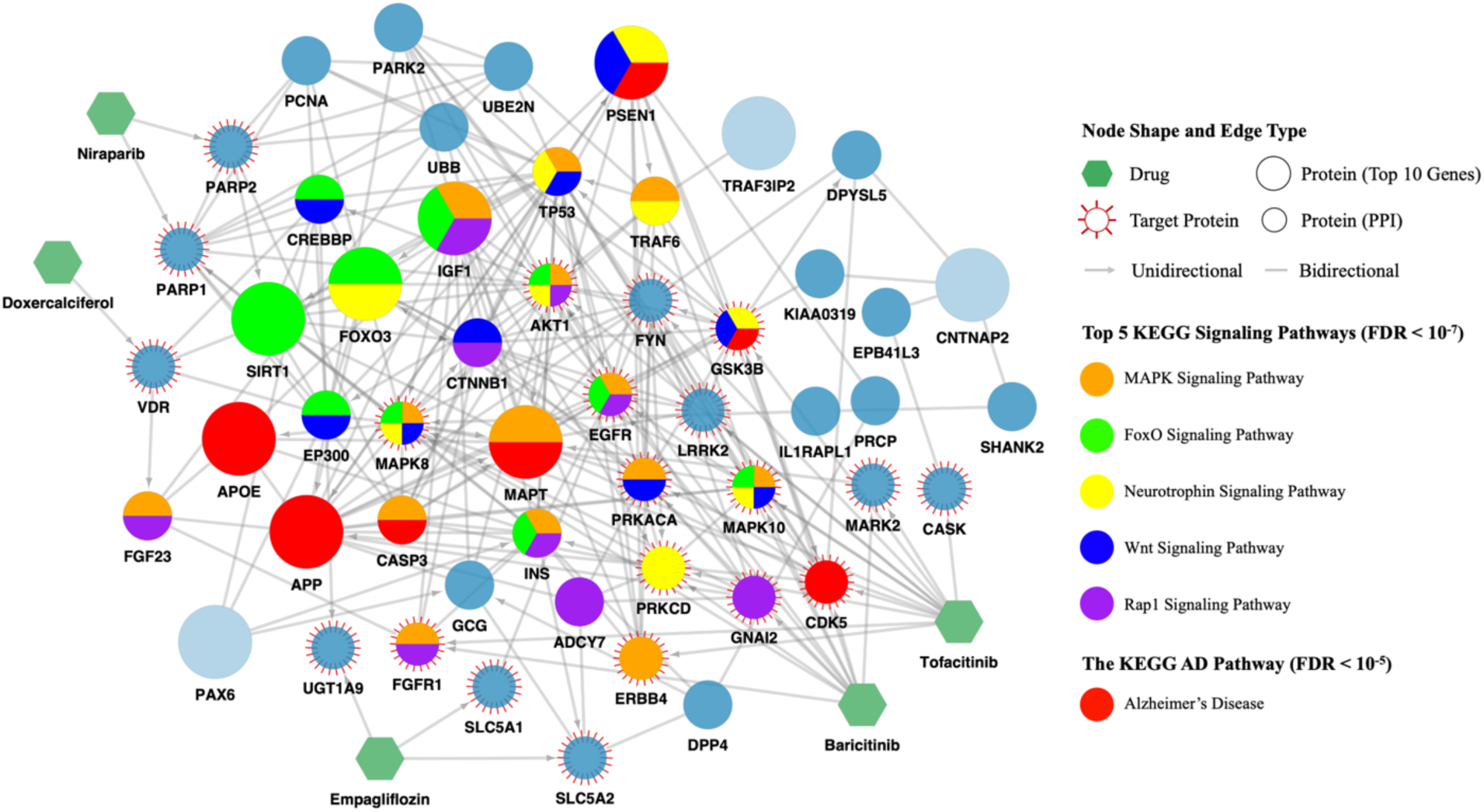
DeepDrug Prediction of Five-drug Combinations and Drug-target-gene Interaction Network

Moreover, using STRING pathway enrichment analysis^72^, Figure 4 reveals the top five KEGG pathways enriched in the PPI subnetwork according to their statistical significance levels. The significance level is measured by the false discovery rate (FDR), and the threshold is set at 5%. The KEGG AD pathway ranks the top 17^th^ among all significant pathways (44 in total), which includes APOE, APP, CASP3, GSK3B, PSEN1, MAPT, and CDK5 in the PPI subnetwork. The top five signaling pathways are all related to AD pathology, covering aging, oxidative stress, neuroinflammation, amyloid beta production, and Tau hyperphosphorylation. Specifically, the MAPK signaling pathway is linked to Tau hyperphosphorylation implicated in brain somatic mutations^73^ and neuroinflammation^74^. The FOXO signaling involves FOXO genes, which are transcription factors that regulate reactive oxygen species (ROS) detoxification, cell cycle/death, and energy/glucose metabolism^75^, and has been linked to longevity, aging^76^, and oxidative stress^77^, with the latter being linked to AD pathogenesis. The neurotrophin signaling pathway is connected to a variety of signaling cascades, including the MAPK and PI3K signaling pathways, which are also linked to Tau hyperphosphorylation^73^ and neuroinflammation^74^. The Wnt signaling pathway inhibits GSK3B, a key kinase involved in Tau phosphorylation, reinforces amyloid beta production via BACE1 activation^78^, and regulates microglial inflammation^79^. In addition to the MAPK and Wnt signaling pathways, the Rap 1 signaling pathway is also considered one of the core neuroinflammation signaling pathways^80^.

Furthermore, Figure 4 reveals how the five drugs in combination can be used to treat AD at the pathway level. Tofacitinib and Baricitinib directly hits all the top five pathways via protein targets such as AKT1, CDK5, EGFR, GSK3B, MAPK8, and MAPK10. In contrast to Tofacitinib and Baricitinib, the other three drugs do not directly target any of the top pathways. The reason why including the other three drugs may be beneficial for AD treatment can be explained in two ways. On the one hand, some of the other three drugs can reinforce Tofacitinib and Baricitinib by indirectly targeting different targets of the same top pathways, thus increasing the synergistic effect of the drug combination^81^. For example, Empagliflozin can target EGFR in the MAPK, FOXO, and Rap1 signaling pathways via SLC5A1. Similarly, Niraparib can target SIRT1 in the FOXO signaling pathway via PARP2, and Doxercalciferol can target FGF23 in the MAPK and Wnt signaling pathways via VDR. The synergistic effect can increase when different targets in the same top pathways crosstalk other AD-associated pathways^81^. For example, in the MAPK signaling pathway, EGFR targeted by Empagliflozin is also involved in the FOXO and the Rap1 signaling pathway, which are linked to neuroinflammation and oxidative stress. On the other hand, some of the other three drugs can serve as complementary drugs by targeting different AD-associated pathways beyond the identified top pathways. For example, to address Tau hyperphosphorylation implicated in different pathways, Doxercalciferol can target the PAX6 signaling pathway^82^ via VDR and its first-order neighbor CREBBP. In parallel, Niraparib can target pathways related to cell cycle and DNA repair in age-related diseases^83^ via PARP1/2 and its first-order neighbor PCNA. According to the parallel pathway theory of drug synergy, which argues that drugs targeting separate pathways to a common biological function are synergistic^84^, these complementary drugs can greatly increase the effectiveness of AD treatment.

## Discussion

Recent developments in AI technologies, such as GNN^85^ and causal AI^86–88^, and the accumulation of large volumes of relevant datasets such as KEGG, DrugBank, CMap, SPIED and DGIdb, have provided new opportunities for an AI-driven drug-repurposing approach to greatly improve the probability of finding effective AD drug candidates, vastly reducing developmental costs and accelerating precision drug identification.

However, three challenges remain to be overcome. Firstly, existing data-driven computational drug-repurposing methods have yet to fully incorporate expert-led domain-specific AD knowledge into a heterogeneous signed directed biomedical graph, including a rich set of nodes and edges, carefully incorporating expert knowledge in graph construction and node/edge weighting to capture the convergence and divergence of crucial pathways associated with AD. Secondly, most existing network-based models determine the efficacy of a repurposed drug by estimating the distance between the drug and disease proteins, based on a simple shortest path distance. Thirdly, the systematic identification of high-order drug combinations to maximize the synergistic effect remains challenging due to high computational costs and the lack of experimental data.

To address these challenges, this study proposes an expert-led AI-driven framework to determine a lead five-drug combination for AD treatments based on a heterogeneous signed directed biomedical graph. Even though the underlying mechanism of AD is less well understood, our proposed GNN model incorporates expert-led domain-specific insights. Such insights include knowledge on long genes, immunological and aging pathways, and somatic mutation markers identified in the blood identified from the previous work^89^ to be closely associated with AD, as well as how to assign weights to the nodes and edges in graph construction. Further, by encoding our biomedical graph via GNN into a new embedding space, DeepDrug can better capture the rich relationship between different nodes in the original graph as compared to what a pure shortest path metric can provide^30^. Nevertheless, with our signed directed edges and with node and edge weight assignments, a more accurate prediction of successful drug candidates can be provided. Finally, by using diminishing return-determined thresholds, we can better select the high-order drug combinations from the top drug candidates to achieve the maximum synergistic combinatory effect for improving the effectiveness of AD treatment.

Capitalizing on the recent findings that long genes are more likely to be affected by somatic mutations^90^, and accumulating somatic mutations are implicated in brain pathology^89^ and AD-risk blood^91^ (see Downey, Lam, Li, and Gozes (2022)^92^ for a detailed review of somatic mutations and AD), we have developed a novel DeepDrug framework and identified the top repurposed five-drug combination candidates. The top 15 drug candidates identified by DeepDrug (see Table 1 in “Results”) are consistent with prior biological and clinical knowledge. Tofacitinib, a top candidate, was previously highlighted in AI-driven AD research and tested in mice models^47,48^. Three of the top five drugs are JAK inhibitors, which can reduce neuroinflammation contributing to AD pathology^49^. For the remaining top drugs, Niraparib, Palbociclib, and Sirolimus can be potential therapeutic agents, given their role in autophagy in AD^50^. Empagliflozin, Febuxostat,

Doxercalciferol, and Zinc Acetate can address AD-associated comorbidities and risk factors (diabetes^52^, gout^53^, vitamin D deficiency^54^, and zinc deficiency^55^). Other promising candidates include Raloxifene for women with AD^56^, Valproic Acid and Olanzapine originally for other neurological diseases but repurposed for AD^57,58^, and Adenosine and Miconazole for reducing neuroinflammation^59,60^.

A lead combination of five drugs, comprising Tofacitinib, Niraparib, Baricitinib, Empagliflozin, and Doxercalciferol, selected from drug combinations of two to six drugs from the top 15 drug candidates, can serve a promising drug combination for AD treatment (see Tables 2 and 3 in “Results”). The selected five-drug combination for AD treatment has significant biological relevance and clinical potential. These drugs, previously approved for treating conditions such as rheumatic diseases, cancer, diabetes, and hyperparathyroidism, are shown to inhibit AD pathology through various mechanisms. Specifically, Tofacitinib and Baricitinib are JAK inhibitors targeting the JAK-STAT pathway, reducing neuroinflammation by inhibiting proinflammatory cytokines mediated by immune cells^49^. Niraparib inhibits PARP enzymes, addressing neurodegeneration by preventing metabolic impairment and regulating cell death^64^. Empagliflozin, an SGLT2 inhibitor, regulates the insulin signaling pathway linking diabetes and AD^65^. Doxercalciferol, a vitamin D analog, protects neurons against inflammation and oxidative stress^66^. The clinical relevance of these drugs in AD treatment is also supported by their involvement in various stages of AD drug discovery, including literature review^50^, epidemiological study^67^, animal models^48,65^, and human clinical trial (Phase II)^68^, highlighting their potential effectiveness in treating AD.

Moreover, t-SNE analysis has revealed the position of the five drugs in the lead combination mapped onto the biomedical embedding space (see Figure 3 in “Results”). On average, this lead combination is closer to AD-associated genes compared to other drug combinations, indicating a higher likelihood of effectively treating AD. These five drugs have been observed in three clusters, suggesting reinforcement and complementary effects. Specifically, Tofacitinib and Baricitinib, both JAK inhibitors, are closely positioned, indicating a strong combinatory effect on neuroinflammation pathways. Niraparib, a PARP inhibitor, and Doxercalciferol, a vitamin D2 analog, are also closely positioned, suggesting anti-inflammatory and oxidative stress reduction through their combined actions. Empagliflozin, an SGLT2 inhibitor to treat diabetes, is distant from the others, indicating its complementary role in targeting the insulin signaling pathway, which are related to glucose, lipid, and energy homeostasis.

Further, network and pathway analysis has provided the mechanism insights into the lead combination of drugs, showing how the drugs and their targets interact with the key AD-associated genes (see Figure 4 in “Results”). Despite not directly targeting the primary genes in the KEGG AD pathway, the selected drug combination can target these genes through PPIs. STRING pathway enrichment analysis has identified the top five KEGG pathways enriched in the PPI subnetwork. These pathways include the MAPK, FOXO, neurotrophin, Wnt, and Rap1 signaling pathways, which are all related to AD mechanisms from different perspectives, including aging, oxidative stress, neuroinflammation, amyloid beta production, and Tau hyperphosphorylation. Tofacitinib and Baricitinib target all top pathways directly, involving key proteins such as AKT1, CDK5, EGFR, GSK3B, MAPK8, and MAPK10. The other three drugs, Empagliflozin, Niraparib, and Doxercalciferol, do not directly target these pathways but reinforce and complement Tofacitinib and Baricitinib. Their reinforcement and complementary effects can be achieved by indirectly targeting the same top pathways or additional AD-related pathways, thus addressing broader aspects of AD pathology and enhancing the synergistic effect. By targeting multiple interconnected pathways, the selected drug combination further reveals its potential efficacy in AD treatment, offering a comprehensive approach to tackling the complex mechanisms of AD.

Based on the calculated drug-gene scores, we have also investigated the rankings of existing approved AD drugs, including Lecanemab and Aducanumab, the new drugs approved by FDA in 2021 and 2023, respectively, though not approved by the EU. Aducanumab has been off the market since 2024^93^. The rankings of the approved AD drugs (out of 503 drugs) in our GNN model are: Memantine (55th), Donepezil (83rd), Galantamine (159th), Rivastigmine (264th), Lecanemab (403rd), and Aducanumab (405th). They do not rank high in our GNN model, most probably because they are targeting at AD symptoms or events that are insufficient in-and-of-themselves to trigger a disease. For example, the monoclonal antibodies, Aducanumab and Lecanemab, target amyloid beta^94^, an upstream biomarker in the AD pathology, which may explain why they are not effective in treating progressed AD. In contrast, the upstream vs. downstream effects of mutations in the genes (such as tau vs. amyloid beta), along with different AD-related pathways implicated in AD brain and blood, have been incorporated into the proposed framework during the biomedical graph construction. The low rankings of approved AD drugs can be attributed to the expert-led knowledge incorporated into the GNN model input (which did not focus on amyloid beta only), such as long genes, co-morbidities, and the importance of AD-specific pathways. The findings from our GNN model are also consistent with the observations from existing AD drug experiments. Until now, no drugs have been proven to treat AD effectively. The repeated failures of the approaches targeting amyloid in AD clinical trials have increasingly pointed to other potential directions in AD drug development, such as the role of neuroinflammation and the immune system^95^. These results, taken together, highlight the importance of incorporating expert-led insights into our proposed AD drug repurposing framework.

Nevertheless, translating AI-driven drug repurposing findings into clinical practice presents significant challenges. Firstly, the limited availability of comprehensive datasets for AD and the lack of effective drugs for its treatment may hinder the training of AI models, potentially leading to biased and inaccurate predictions. Secondly, the expert-led biomedical graph structure simplifies complex biological systems, possibly resulting in an inadequate representation of drug combinations, particularly at the molecular level. Thirdly, AI-driven repurposing models operate as “black boxes”, making it difficult to understand the rationale behind their drug prediction processes. This lack of interpretability may impede the adoption of AI-driven drug-repurposing in clinical settings. Finally, validating AI-driven findings in rigorous experimental and clinical studies is often costly and time-consuming, requiring substantial effort and resource.

To tackle these challenges, our current AI-driven GNN model can be improved further in the following directions: Firstly, given that no effective drug is yet available for AD, to facilitate the GNN model’s learning towards which drug can “treat” which gene, multi-task transfer learning can be used to simultaneously repurpose drugs for AD and other related diseases (where ground truth drugs may be available). Secondly, the GNN model structure can utilize hierarchical graph neural networks^96,97^ to better represent drug combinations at the molecular level (e.g., a drug consisting of atoms and bonds^98^ and a drug combination consisting of multiple drugs) while capturing the relationships of nodes at different layers through graph attention mechanisms. The fine-grained molecular representation can also improve the calculation of suitable drug doses to prevent adverse effects^99^, and facilitate the prediction of blood-brain barrier permeability^100^, making it possible to deliver effective drugs to treat AD brains. Thirdly, the interpretability of the GNN model can be improved. Instead of starting with the 310 genes associated with AD in our biomedical graph, we may start with the genes most causal to AD. These genes may be determined as the most upstream genes in AD-associated pathways. In addition to the pathway enrichment analysis, the contribution of local subgraphs^101^, in particular, genetic pathways, in calculating the drug-gene score can also be investigated to better understand the importance of different pathways in AD drug-repurposing. Such interpretations from a “black box”, combined with expert insights, will promote the feedback loop in the proposed expert-led AI-driven approach.

Finally, having identified the lead combination of drugs *in silico*, the last step - testing of drug efficacy *in vitro* and *in vivo,* remains to be undertaken. In the future, our DeepDrug framework can be applied to repurpose drugs for other neuro-degenerative and neuro- developmental diseases, such as Parkinson’s disease, autism spectrum disorders, PTSD, multiple sclerosis, and schizophrenia.

## Methods

DeepDrug, an AD drug-repurposing framework, consists of a three-stage development. First, a biomedical graph was created, linking genes, proteins, drug targets, and drugs to capture the network characteristics of AD pathology (see “Heterogeneous Signed Directed Biomedical Graph Construction”). Second, a GNN-based framework was developed to learn the biomedical graph structure and represent drug and gene nodes as lower-dimensional vectors (see “Graph Neural Network to Embed Drug and Gene Nodes” and “GNN Model Training and Evaluation”). Third, drug scoring and selection were performed to identify the top drug candidates and a lead drug combination (see “Scoring and Selection of Drug and Drug Combination” and “Drug Combination Optimisation”).

Figure 5 provides an overview of DeepDrug, an AD drug-repurposing framework, from two complementary perspectives, namely, a domain-specific knowledge-inspired approach, and an AI-driven approach. On the left side of Figure 5, the domain-specific knowledge-inspired approach instructs the construction of a heterogeneous signed directed biomedical graph. The biomedical graph consists of four types of nodes, including genes, proteins, targets, and drugs, and four types of edges between nodes, including gene-protein, protein-protein, drug-target, and drug-drug edges. It starts from identifying key genes implicated in AD and approved drugs relevant to AD. Protein targets bound to the selected drugs are connected to the proteins corresponding to the identified genes through protein-protein interactions (PPIs). The identified genes also undergo pathway enrichment analysis to determine their roles in AD-associated pathways in the PPI network. Finally, expert-guided knowledge is integrated into the biomedical graph by determining edge directions (e.g., whether one protein acts on another protein) and signs (e.g., whether a drug activates or inhibits its target) and assigning weights to nodes and edges (e.g., gene nodes weighted by their importance to AD). The details regarding how domain-specific knowledge is integrated into the biomedical graph are elaborated in “Heterogeneous Signed Directed Biomedical Graph Construction”.

**Figure 5.**
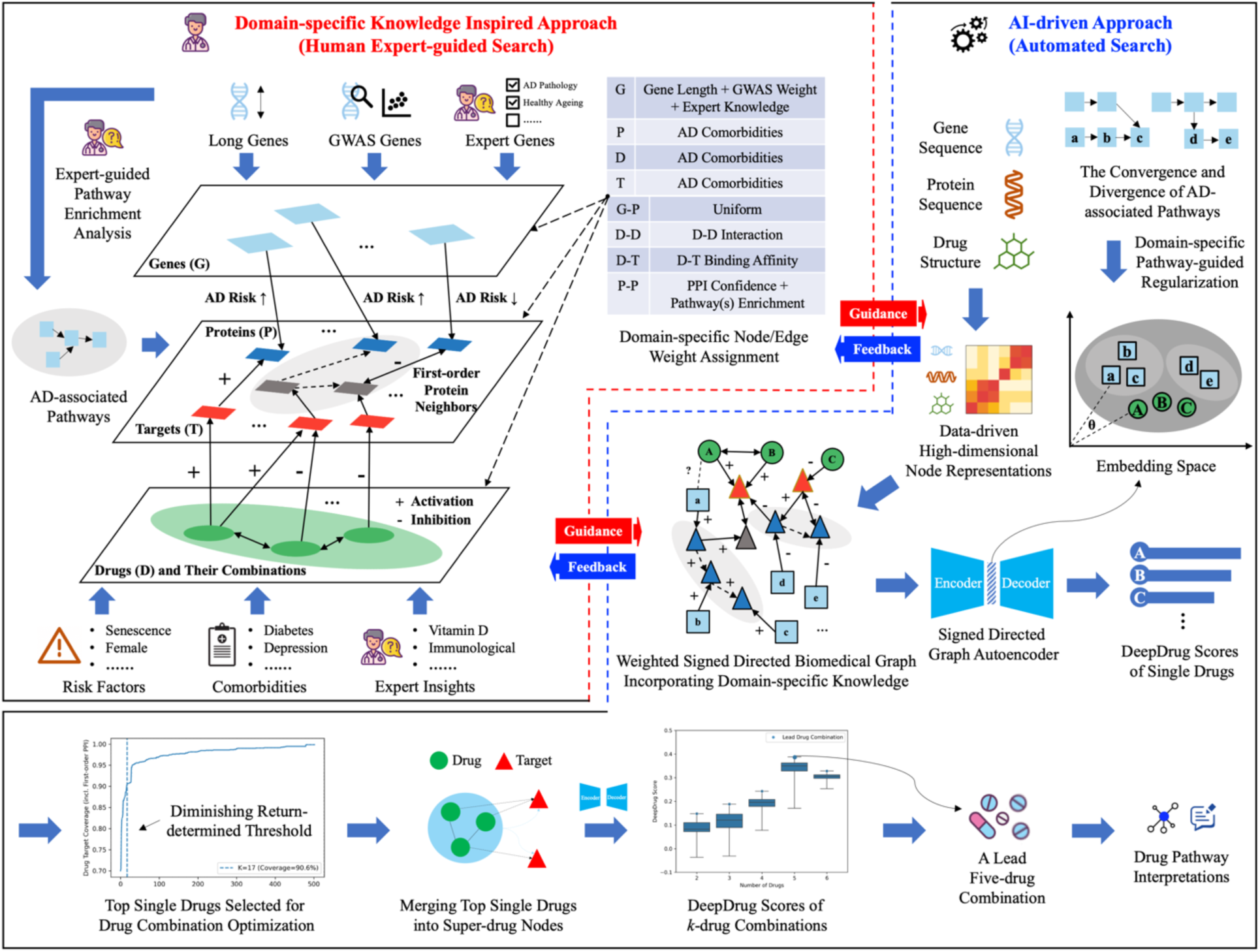
Expert-guided AI-driven DeepDrug Framework

On the right side of Figure 5, the AI-driven approach leverages on the expert-guided biomedical graph plus the rich representation of the nodes (gene sequences, protein sequences, and drug structures), mapping them onto a lower-dimensional space (see “Graph Neural Network to Embed Drug and Gene Nodes”). The AI model is trained to understand complex relationships within the biomedical graph, using domain-specific pathway-guided regularization to improve biological relevance in the lower-dimensional space (see “GNN Model Training and Evaluation”). Furthermore, the iterative feedback loop between the two approaches can improve the accuracy and reliability of the proposed framework. This dynamic collaboration between human expertise and AI involves incorporating expert insights into the initial setup and continuous adjustments of the AI methods, ensuring that the automated processes are consistently refined and aligned with the latest domain knowledge.

### Heterogeneous Signed Directed Biomedical Graph Construction

A heterogeneous signed directed biomedical graph was constructed, consisting of four types of nodes, including genes, proteins, drugs, and drug targets, and four types of edges between nodes, including gene-protein, protein-protein, drug-target, and drug-drug edges. After graph construction, isolated nodes were removed from the graph. The detailed graph construction procedure, including how domain-specific knowledge was incorporated into the biomedical graph, can be found as follows.

*Gene Nodes:* Two types of gene mutations relevant to AD, namely, the somatic mutations and the germline mutations, were considered. For the somatic mutations, their gene nodes were obtained from the long genes^90^ and somatic mutation markers identified in the blood^91^. For the germline mutations, their gene nodes were obtained from the high-risk genes identified from GWAS meta-analysis data^102^. Other important genes relevant to (i) AD pathology and (ii) healthy aging were also selected from expert-led insights. The gene nodes were weighted according to their chance of mutation and importance to AD progression (see Table S1 in Supplementary Data for the selected genes and their weights).

Specifically, data on the somatic mutations implicated in the AD pathology was obtained from Soheili-Nezhad et al. (2021)^90^, constituting a total of 272 long genes. GWAS data was obtained from the Phase 3 summary statistics in Jansen et al. (2019)^102^, for a total of 30 significant genes. 10 expert-led genes were chosen based on the expert knowledge of our neuroscience co-authors, who recently discovered ADNP and SHANK3 mutations as the key somatic mutation markers of autism linked to AD^103^. In addition, her team has proven that critical somatic mutations in ADNP correlate with AD Tau pathology^89^, and verified them as driving an early AD-like Tau pathology in a genome-edited (CRISPR/Cas9) mouse model^104^. In total, 310 genes were selected. Only two overlaps between the three sets of genes could be identified, including APOE, between GWAS and the expert-led, and CNTNAP2, between the long gene set and GWAS (see Figure 6).

**Figure 6.**
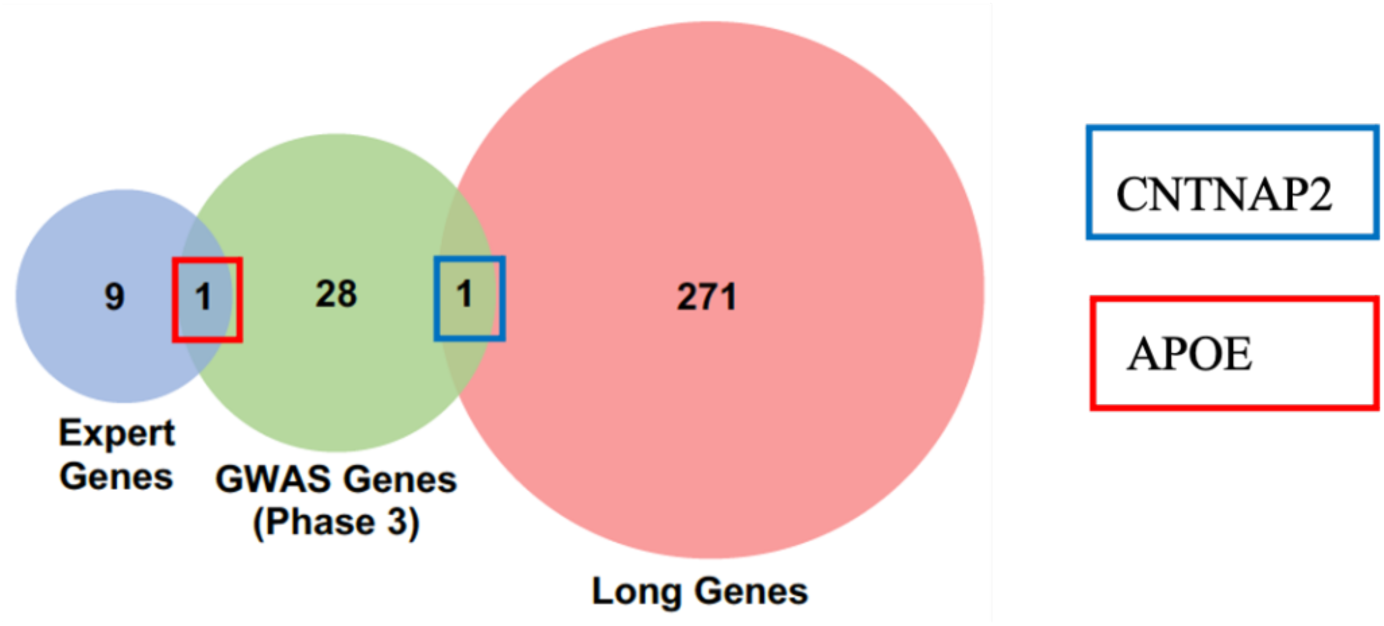
The 310 Gene Nodes Incorporating Expert-led Knowledge.

The weights of the 310 gene nodes (a total of three sets of genes) were calculated based on their lengths and normalized to [1, 2]. For all GWAS genes, their normalized -log(p-value), normalized to [1, 2], was added to their existing normalized weight based on gene length. For all expert-led genes, their existing weights (based on normalized gene length plus additional normalized weights from the GWAS step) were increased by 1, to reflect the importance of these genes. Finally, all the gene nodes were renormalized to [1, 2].

In addition to the node weights guided by AD domain-specific knowledge, a high-dimensional weight vector was automatically generated for each gene node based on the corresponding protein-coding sequencing data. Similar to *n*-grams and bag-of-words used in natural language processing tasks, the *k*-mer method was used to (i) decompose each mRNA sequence into a sequence of all possible *k* nucleotides, (ii) calculate the number of times that each *k*-mer appeared in each sequence, and (iii) convert each sequence into a count vector. Finally, the singular value decomposition (SVD) method was used to compress each gene vector into a fixed-length vector.

*Drug Nodes:* Based on the approved drugs (including drugs approved by the FDA or other drug regulatory authorities outside the US), drug nodes were selected using the top co-morbidities and risk factors related to AD as well as expert-led insights. The drug nodes were weighted according to AD co-morbidities (see Table S2 in Supplementary Data for the list of drugs identified and their weights).

Specifically, data on the drugs were obtained from DrugBank (version 5.1.10)^105^, an online database containing information on drugs, drug targets, and drug-drug interactions. The keywords used to find the relevant drugs based on drug description, indication, pharmacodynamics, and mechanism-of-action were (i) the top two risk factors of AD: ‘senescence’, ‘female’, (ii) the top four co-morbidities of AD: ‘Diabetes’, ‘Cardiovascular disease’, ‘Depression’, ‘Inflammatory bowel disease’, (iii) miscellaneous terms guided by expert insights: ‘Reactive oxygen species’, ‘Vitamin D’, ‘immunological’, ‘autoimmune’, and (iv) the disease itself: ‘dementia’ (given that AD is the most common form of dementia) and ‘Alzheimer’s’. After (i) filtering out the vaccines, nutrition supplements, and agents for medical imaging, (ii) selecting drugs whose target organism is human or not specified, and (iii) choosing only the drugs that are approved and not withdrawn or prohibited, the total number of drug candidates was 503.

The drug nodes were weighted by the odds ratio of the key comorbidities they can treat. Lin et al. (2018)^106^ reported the odds ratio of the three co-morbidities of ‘diabetes’, ‘cardiovascular disease’, and ‘depression’ in various combinations. This study, with 49747 subjects aged ≥65 years in 2000, was based on the Taiwan National Health Insurance Research Database, with subjects stratified by the presence and absence of dementia from 2000 to 2010. Dementia subjects (4749 out of the total cohorts) were those first diagnosed with dementia from 2000 to 2009. Table 4 shows the weights assigned to drugs that treat various combinations of these three comorbidities. The rationale is that, if a drug treats the key co-morbidities of dementia, the most prevalent form of AD, it shall top the search list and be accorded a higher weighting subsequently. Meanwhile, the weight of a combination of co-morbidities may not be equivalent to the sum of the individual co-morbidities. In addition, the weight of a single co-morbidity can be higher than a combination including this co-morbidity. Based on the weighting rationale, the weights of the drug nodes were normalized to [1, 2].

**Table 4.** Weights Assigned to Drug Nodes Based on Different Combinations of Comorbidities Being Treated.

| Combination of Co-morbidities Treated | Weights Assigned (Based on Odds Ratio obtained from Lin et al. (2018) <sup>106</sup>) |
| --- | --- |
| Depression | 4.938 |
| Vascular Disease | 3.261 |
| Diabetes | 2.321 |
| Depression + Vascular Disease | 6.726 |
| Depression + Diabetes | 4.787 |
| Vascular Disease + Diabetes | 3.161 |
| Depression + Vascular Disease + Diabetes | 6.521 |

In addition to the expert-guided node weights, a high-dimensional weight vector was automatically generated for each drug node, based on the corresponding chemical structure data. Drugs were small molecules or proteins (antibodies) in this study. For small molecules, a fingerprint was generated for each drug compound using RDKit^107^. For protein/peptide drugs, their weight vectors were calculated along with the protein nodes (see below). Finally, the SVD method was used to compress each drug vector into a fixed-length vector.

*Target Nodes:* Target nodes were obtained along with the drug nodes, as each drug has one or more specific targets. The target nodes were weighted according to AD co-morbidities. (see Table S3 in Supplementary Data for the list of protein targets identified and their weights)

Specifically, drug targets were obtained from four major biochemical databases, including DrugBank (version 5.1.10), DrugCentral (as of Aug 2022), ChEMBL (version 31), and BindingDB (version 2023m0). For targets derived from DrugBank, four target types were provided, including targets, enzymes, carriers, and transporters. Targets and enzymes were selected, while carriers and transporters were discarded. All drug targets were proteins in this study. Drug targets from different databases were combined, resulting in 1,525 drug targets in total.

The target nodes were weighted by their involvement in the top three co-morbidities of AD and were later normalized to [1, 2] along with the other proteins (see below).

In addition to the expert-guided node weights, a high-dimensional weight vector was automatically generated for each target node based on the corresponding protein sequencing data (see below).

*Protein Nodes:* Protein nodes were initially obtained according to the gene and the target nodes, as each gene had an equivalent protein and all drug targets were proteins in this study. Next, the first-order neighbours of the initial protein nodes were included in the graph according to the human protein-protein interaction (PPI) data. The protein nodes were weighted by the same method as that of the target nodes (see Table S3 in Supplementary Data for the list of proteins and their weights).

Specifically, data on the proteins were obtained from STRING (version 11.0), an online curated biological database integrating different sources of evidence and providing a combined score (ranging from zero to one) to indicate whether a PPI is biologically meaningful ^72^. The data were first filtered to include only the proteins in human beings. Next, using the 310 proteins arising from the 310 genes and the 1,525 protein targets resulted in 1,804 proteins. Next, the 1,804 proteins’ first-order neighbours, with high PPI confidence of more than 0.7, were added, giving a total of 11,222 proteins.

The protein nodes were weighted by their involvement in the top three co-morbidities. The weights were then normalized to [1, 2] along with the target nodes.

In addition to the expert-guided node weights, a high-dimensional weight vector was automatically generated for each protein/target node and protein/peptide drug node, based on the corresponding amino acid sequencing data. The *k*-mer method was used to (i) decompose each protein sequence into a sequence of all possible *k* amino acids, (ii) count the number of times that each *k*-mer appeared in each sequence, and (iii) convert each sequence into a count vector. Finally, the SVD method was used to compress each protein vector into a fixed-length vector.

Next, we determined the edges between the nodes and the associated weights. There were four types of edges, namely, gene-protein, drug-drug, drug-target, and protein-protein. All edges were directed, and some were signed. Gene-protein and drug-target edges were assumed to be signed and unidirectional. A positive or negative gene-protein edge indicated that mutations in the gene and resulting protein dysfunction would be positively or negatively associated with AD. A positive or negative drug-target edge indicated that the drug would activate or inhibit its binding protein. Protein-protein edges were considered bidirectional by default, except that some were unidirectional according to the PPI data. Similarly, protein-protein edges were considered unsigned by default, except that protein action types were specified in the PPI data. A positive or negative protein-protein edge represented activation or inhibition, respectively. All drug-drug edges were assumed to be unsigned and bidirectional to account for the magnitude of side effects when two drugs are taken together. Detailed descriptions of how the signs, directions, and weights of these edges were determined can be found below, including the list and the signs/directions of these edges, plus their associated weights (see Tables S4-7 in Supplementary Data).

*Gene-Protein Edges:* For the gene-protein edges, a unidirectional relationship between the genes and the proteins was established directly, as one gene is matched to one single protein. The gene-protein edges were weighted uniformly. It can be further weighted when the mRNA stability data is available (e.g., how many proteins can one mRNA create before degrading). The sign of the gene-protein edges was determined based on how mutations in the genes were linked to AD.

With the 310 gene nodes, 310 gene-protein edges could be identified. Due to a lack of mRNA stability data, the gene-protein edges were weighted uniformly with a default average weight of 1.5 (after normalizing to [1, 2]). For the 272 long genes, the sign was set to positive (+1), given that somatic mutations in these long genes were positively linked to AD by showing reduced gene expression and inhibited synaptic-related pathways in AD brains. For the GWAS genes, the sign was determined by the corresponding *Z* score – a negative *Z* score suggested a negative link between the GWAS gene and AD risk, resulting in a negative sign (-1), and vice versa. For the expert-led AD-risk genes, the sign was set to positive (+1).

*Drug-Drug Edges:* A default bidirectional relationship was established for the edges between the drugs. The drug-drug edges were weighted by text-mining the drug-drug interaction documents (i.e., whether two drugs increased or decreased risk). The drug-drug edges were unsigned given that only the magnitude of side effects was considered.

Drug-drug interaction data were obtained from the DrugBank database (version 5.1.10)^105^. A total of 36,532 bidirectional drug-drug edges were established. The edges were weighted using the text information of available detailed drug-drug interactions from DrugBank. Specifically, a default weight of 1.5 was given, and, through text-mining for the words ‘increase risk’ or ‘decrease risk’, if the interaction between two drugs was positive (reduced risk), 1 was added to the weight, else 1 was subtracted from the weight (increased risk). The weights of the drug edges were then normalized to [1, 2]. The sign of drug-drug edges was set to 0.

*Drug-Target Edges:* A unidirectional relationship between drugs and targets was established directly, as each drug has its own (one or more) specific targets. The signs and weights of drug-target edges were determined by the drug-target binding affinity data.

Specifically, the drug-target interactions from DrugBank (version 5.1.10)^105^, DrugCentral (as of August 2022)^108^, ChEMBL (version 31)^109^, and BindingDB (version 2023m0)^110^ were obtained and combined, resulting in 7,379 drug-target edges. The corresponding drug-target affinity measures, including IC_50_, EC_50_, K_i_, and K_d_, were available from DrugCentral, ChEMBL, and BindingDB. For all affinity metrics, the unit was first normalized to nanomolar (nM). Repeated measurements from different data sources and experiments were then aggregated based on the median values to reduce the impacts of outliers. The sign of drug-target edges was determined by the drug-target action type. A positive sign (+1) was given if the drug-target action had keywords including “activator”, “agonist”, “opener”, or “positive”, while a negative sign (-1) was given if the drug-target action had keywords including “inhibitor”, “antagonist”, “blocker”, or “negative”. For a negative drug-target relationship, the drug affinity score was calculated based on the KIBA score (an integrated inhibition metric based on IC_50_, K_i_, and K_d_^111^). For a positive drug-target relationship, the drug affinity score was K_d_ or EC_50_, depending on which metric was available. Drug-target weights were then calculated using a -log(x) transformation on the affinity scores^107^.

Missing drug-target signs and weights were predicted by the high-dimensional drug and target weight vectors using gradient-boosting decision trees, with the assumption that the drug affinity measures between similar drugs and targets (measured by their structure information) are related. Finally, the weights of the drug-target edges were normalized to [1, 2]. A total of 7,379 drug-target edges were identified.

*Protein-Protein Edges:* A mixture of unidirectional and bidirectional relationships was established between proteins. The signs and weights of protein-protein edges were determined by the PPI data. Specifically, the direction of protein-protein interaction was set to bidirectional by default, except that the directionality was indicated (i.e., one protein is acting on another protein) in the STRING database. The protein-protein edges were weighted by the confidence of the two involved proteins, a combined score integrating evidence from different data sources, without being normalized. The protein-protein edges were unsigned by default (the sign was set to 0). The sign was further determined by the protein-protein action type if such information was available. A positive sign (+1) was adopted if the action type was “activation”, while a negative sign (-1) was adopted if the action type was “inhibition”. A total of 243,413 protein-protein edges were established.

*Pathways:* Based on the expert-led domain-specific knowledge, the weights of some protein-protein edges were further increased according to the enrichment value of certain AD-associated pathways if they were part of those pathways. This also covers the overlapping pathways, whilst the protein-protein edges present in multiple pathways were given the additional weight for each pathway, with the assumption that these edges can be considered to be the common denominators of AD.

Specifically, all genes (long, GWAS, and expert-led genes) were used to generate the AD-related pathways, using the STRING pathway analysis^72^. A total of 16 statistically significant pathways, with a false discovery rate (FDR) less than 0.05, were obtained from the KEGG, Reactome, and WiKiPathways databases. The log(-FDR) values were normalized to [1, 2]. Next, for those not-yet-normalized PPI edges which contained proteins involved in one/some significant pathway(s), their weights were further added by the normalized log(-FDR) value of the corresponding pathway(s). Finally, the protein-protein edge weights were renormalized to [1, 2].

Figure 7 shows the DeepDrug directed biomedical graph. This directed biomedical graph consists of the four node types and their edges (interactions). This is a simplified version including the largest connected component of the biomedical graph consisting of only the top 50 nodes of each node type. This illustrates how the AD-relevant genes are connected to the proteins, then to the drug targets, and finally to the drugs.

**Figure 7.**
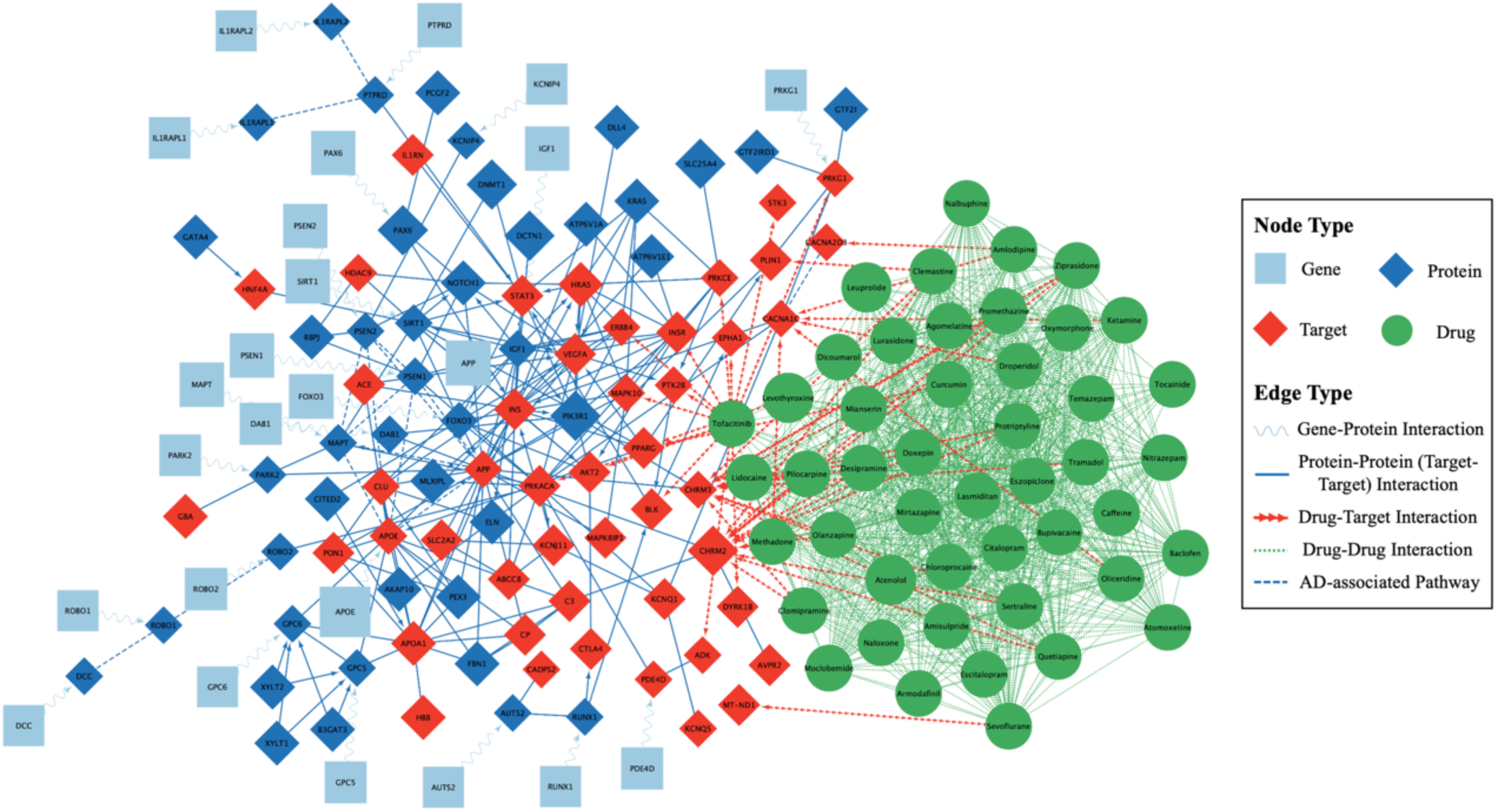
DeepDrug Directed Biomedical Graph (Simplified Version)

### Graph Neural Network to Embed Drug and Gene Nodes

A signed directed GNN framework was developed to map all nodes in the heterogeneous biomedical graph, including gene and drug nodes, to the same embedding space. At a high level, a graph autoencoder (GAE) framework^112^ was adopted to learn a low-dimensional representation of each node based on the node features and a signed directed graph adjacency matrix measuring how nodes are inter-connected (see Figure 8). The novelties of the proposed GNN framework are four-fold (see Figure 8). Firstly, expert-guided weights and data-driven high-dimensional features from the biological structure information are incorporated into node features. Secondly, expert-guided weights, positive/negative edges, and directed edges are incorporated into the graph adjacency matrix. Thirdly, a signed directed GAE is used to embed gene and drug nodes while accounting for positive/negative edges and directed edges. Fourthly, domain-specific pathway-guided regularization is adopted to align gene nodes along the same pathway in the embedding space.

**Figure 8.**
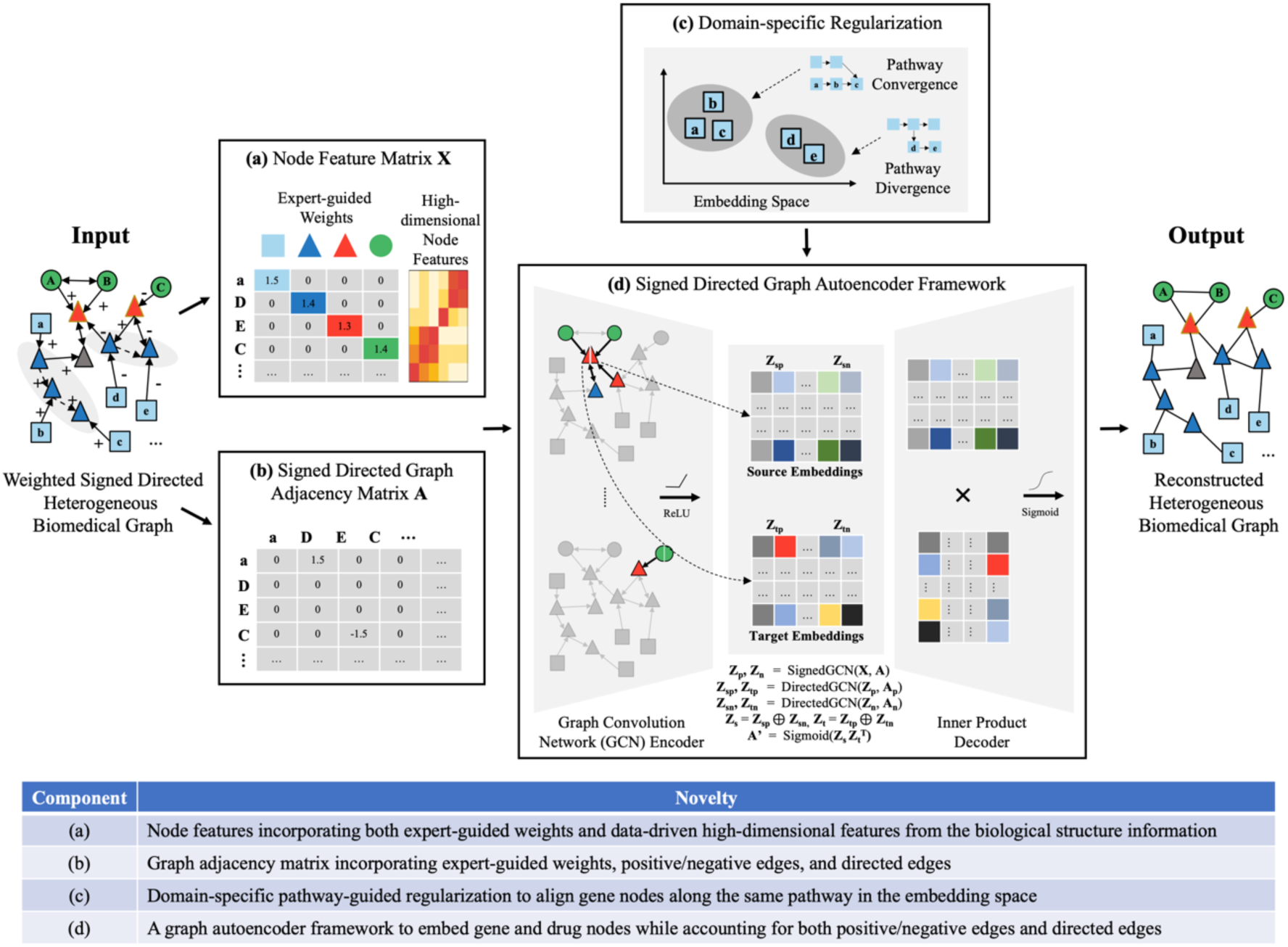
Graph Neural Network to Embed Drug and Gene Nodes and Its Novelties

Specifically, the signed directed GAE consists of two parts, namely, an encoder and a decoder. The encoder projects all nodes to a low-dimensional embedding space using signed and directed GCN layers based on the node features and edge weights while the decoder performs an inner product operation on the node embeddings to reconstruct the graph. The GAE framework can be used for unsupervised graph representation learning without ground truths from downstream tasks (i.e., which drugs can treat AD).

An adjacency matrix ***A****^N^*^×*N*^, which represents how nodes are connected and how edges are weighted, is obtained from the multimodal graph ***G*** consisting of *N* nodes. The adjacency matrix is further converted to two matrices 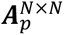 and 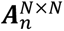 accounting for positive edges and negative edges. Unsigned edges are incorporated into both matrices. An expert-guided node feature matrix 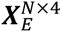 is obtained from ***G***, where each node is represented by a four-dimension feature vector. The four-dimension vector is the product of a node’s weight and a one-hot vector representing the node’s type (e.g., [1, 0, 0, 0] for a gene node). In addition, a data-driven node feature matrix 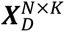 is obtained from ***G***, where each node is represented by a *K*-dimension feature vector calculated from its biological structure information. The final node feature matrix ***X****^N^*^×*M*^ is the concatenation of 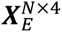 and 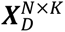. The encoder first takes ***A****^N^*^×*N*^ and ***X****^N^*^×*M*^ as the input to produce positive and negative embeddings for each node via a signed GCN^113^. The positive and negative embeddings are then used to generate source and target embeddings via directed GCNs^114^. The decoder model optimizes low-dimensional node embeddings (accounting for edge sign and direction) to reconstruct the network structure. Conceptually, the GNN framework is described as follows:

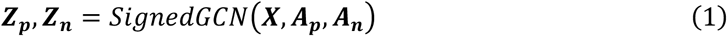

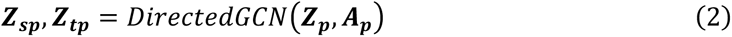

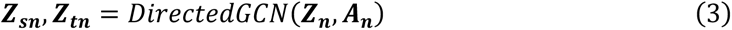

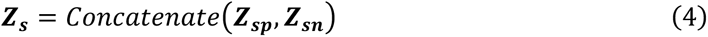

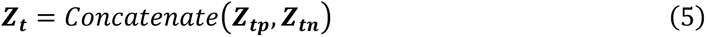

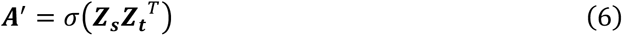

where

SignedGCN is a signed GCN with the ReLU activation function,

DirectedGCN is a directed GCN with the ReLU activation function,

***Z*** represents low-dimensional node embeddings,

*σ* is the sigmoid activation function,

and ***A***′ is the edge probability matrix of all node pairs.

### GNN Model Training and Evaluation

After the biomedical graph construction, the GNN model was trained through a link prediction task on the biomedical graph to optimize its parameters. The task is a binary classification problem, which predicts the closeness between two nodes on the graph, thus allowing one to indicate which drugs are candidates for a certain gene node. Furthermore, to account for the convergence and divergence of AD-associated pathways, a pathway-guided regularization term is incorporated into the training process to force the gene nodes along the same pathway to stay closer to each other in the embedding space, while preserving the convergence and divergence information. Specifically, the regularization term consists of two classification tasks based on node embeddings: a cross-entropy loss for node type classification (one node must belong to one of the four node types) and a binary cross-entropy loss for pathway classification (one node can be in zero, one, or more pathways). The detailed training procedure is shown in Algorithm 1.

#### Algorithm 1. Model Training

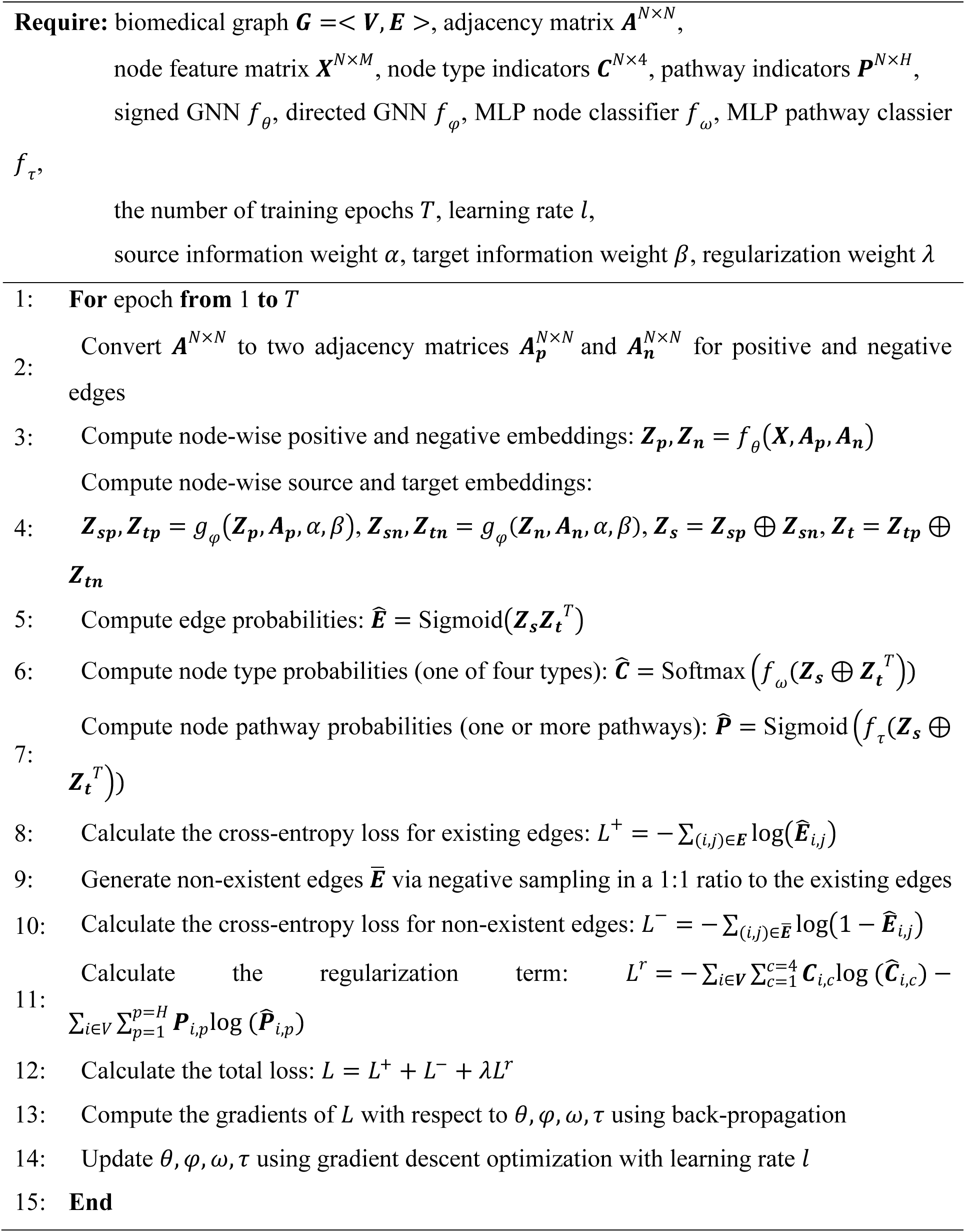
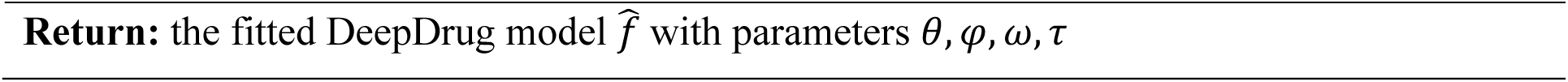

The hyperparameter settings are listed as follows. The dimension of node feature vectors was 260, including four dimensions from the expert-guided node weight vector (representing gene, protein, drug, and target nodes) and 256 dimensions derived from the corresponding biological structure (gene sequence, protein sequence, or drug compound). Each GCN in the encoder had two layers, and the hidden unit size was 128. The embedding dimension size was selected empirically and set to 32. The number of training epochs *T* was 100. The Adam optimizer was used to learn the model parameters, with a learning rate *l* of 0.01. For the directed GCNs, two hyperparameters α and β were used ([0.4, 0.6], [0.5, 0.5], or [0.6, 0.4]) to control the relative importance of source and target information. The weight of the pathway-guided regularization term λ was 0.1, 0.5, or 1.0.

We performed 10-fold cross-validation for model training and evaluation. Specifically, the whole dataset (i.e., all existing edges on the biomedical graph) was randomly split into ten folds of equal size. We held out one of the ten folds (i.e., 10% of all samples) as the testing set. We used a 90/10 split of the remaining data as the training and validation sets. The validation set was used for hyperparameter selection and early stopping during model training. This procedure was repeated ten times, resulting in ten evaluations of the model performance, one for each data fold, allowing for held-out testing on every sample and improving the robustness of model evaluation.

During cross-validation, the trained model was used to predict the existence of edges from the testing set based on the optimized node embeddings (see Equation (6)). This link prediction task is a binary classification problem, where positive and negative labels represent the existence and non-existent edges, respectively. The Area Under the ROC Curve (AUC) and Average Precision (AP), which are two commonly used evaluation metrics for link prediction^115^, were selected for model evaluation. AUC is the area under the receiver operating characteristic (ROC) curve, summarizing the true positive rate (sensitivity or recall) and the false positive rate (1- specificity) at various thresholds into a single value. AUC ranges from 0 to 1, with an AUC of 1 indicating perfect performance. AP is the area under the precision-recall curve, summarizing the trade-off between precision and recall at various thresholds into a single value. AP ranges from 0 to 1, with an AP of 1 indicating perfect performance.

The mean AUC and AP scores and their standard deviations were calculated across all folds (see Figure 9). Results have shown that the signed directed GNN model can achieve more than 97% classification accuracy on average (AUC=0.977 and AP=0.979). The classification performance is consistent across different folds, suggesting the robustness of the proposed model in predicting edges on the biomedical graph and capturing the underlying representations of the biomedical graph structure.

**Figure 9.**
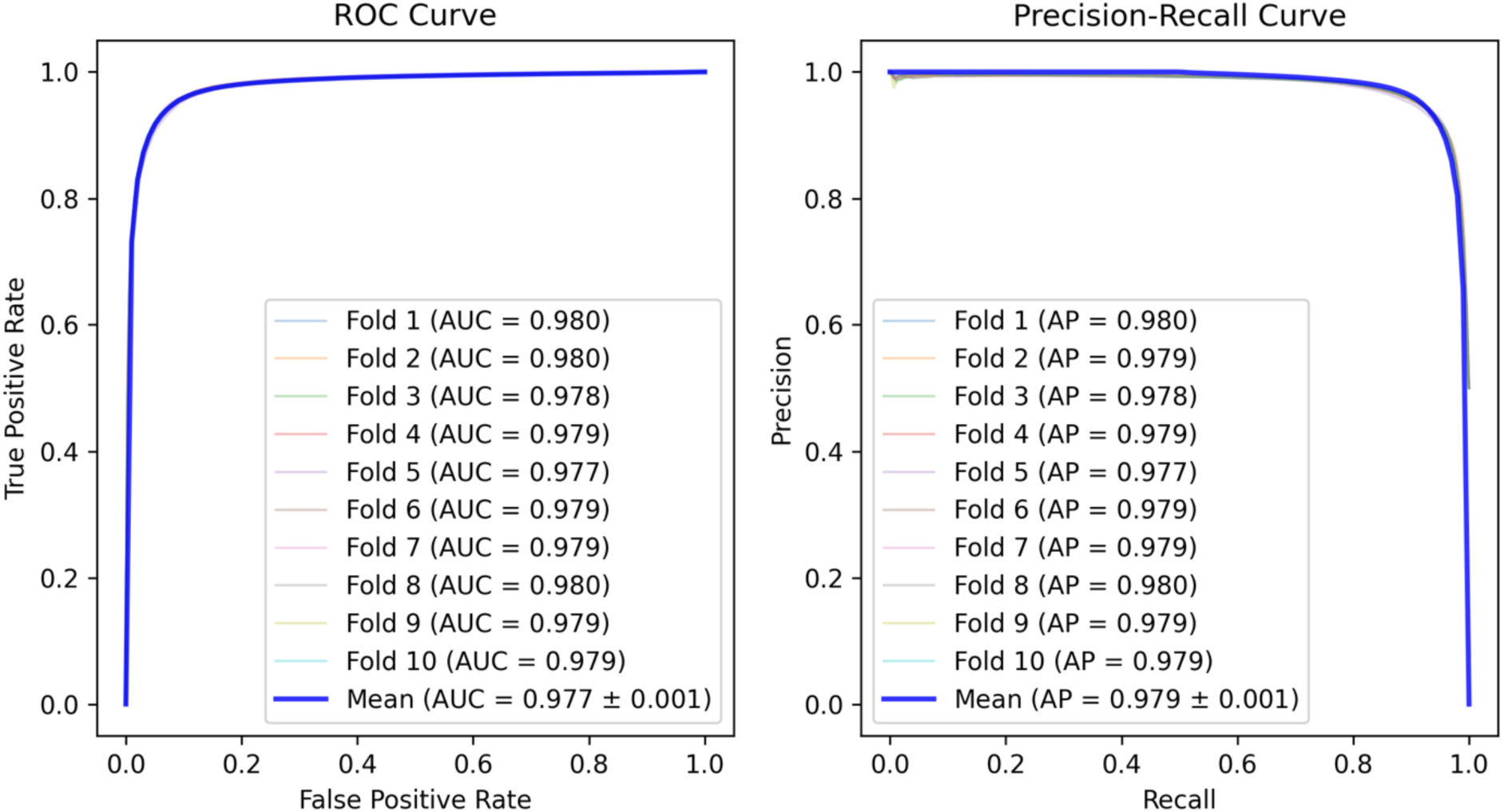
ROC and Precision-recall Curves of the GNN Model

### Scoring and Selection of Drug and Drug Combination

After cross-validation, we calculated drug scores and selected the top drug candidates based on the ten models trained on all folds. The drug scoring was performed in two steps. First, all data were fed into each trained model to obtain an embedding vector for each drug and gene node. We calculated the drug-gene score 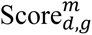 for each drug *d* and gene *g* using model *m*. Specifically, the closer a drug is to the AD-associated gene nodes, the more likely it is a higher-ranking AD drug candidate. Based on the signs of gene-protein edges and drug-target edges, we first calculated the drug-gene scores based on the anti-correlation between the drug embedding and the gene embedding, with the aim to find drug inhibitors close to the AD-risk genes or drug activators close to the AD-protective genes (see Equation (7)).

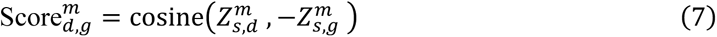

where

*d* represents one drug,

*g* represents one gene,

and 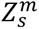 is the node’s embedding vector accounting for the drug-target direction or gene-protein direction using the model trained on fold *m*.

Next, we calculated the average DeepDrug score of each single drug candidate Score_L_ based on the average drug-gene score across all AD-associated genes and all trained models (see Equation (8)).

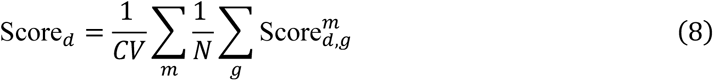

where

*d* represents one drug,

*g* represents one gene,

*m* represents the model trained on fold *m*,

*N* is the total number AD-associated genes,

and *CV* is the total number of cross-validation folds.

Further, we selected the top *K* single drug candidates. The top *K* single drug candidates were used to construct 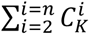 drug combinations for optimisation. *K* was chosen to select complementary drugs as much as possible (in terms of the coverage of proteins they could target) while accounting for the combinatorial cost. The intuition was that the marginal benefit of including more drugs to cover more drug targets would decrease at a certain point. Specifically, the optimal *K* was determined by a drug-target coverage curve, an increasing function characterizing the trade-off relationship between the benefit and the cost. The benefit was the number of targets to be covered, including the direct targets via drug-target interactions and indirect targets through the first-order PPI. The cost was the number of drug candidates to be incorporated into drug combinations. The number of combinations was an exponentially large number, e.g., 1,225 combinations when choosing two out of 50 drugs and more than 15 million combinations when choosing six out of 50 drugs.

### Drug Combination Optimisation

Based on the trade-offs between deriving a higher drug-target coverage and a minimum number of drug candidates, the top 15 drug candidates, covering more than 90% of all targets, were chosen based on the cut-off point of diminishing returns. The cut-off point refers to the point when a high coverage of drug targets has been reached to capture the complementary effects of different drugs, while the number of drug candidates selected is minimal to reduce the computational cost of drug combination optimization (see Figure 10).

**Figure 10.**
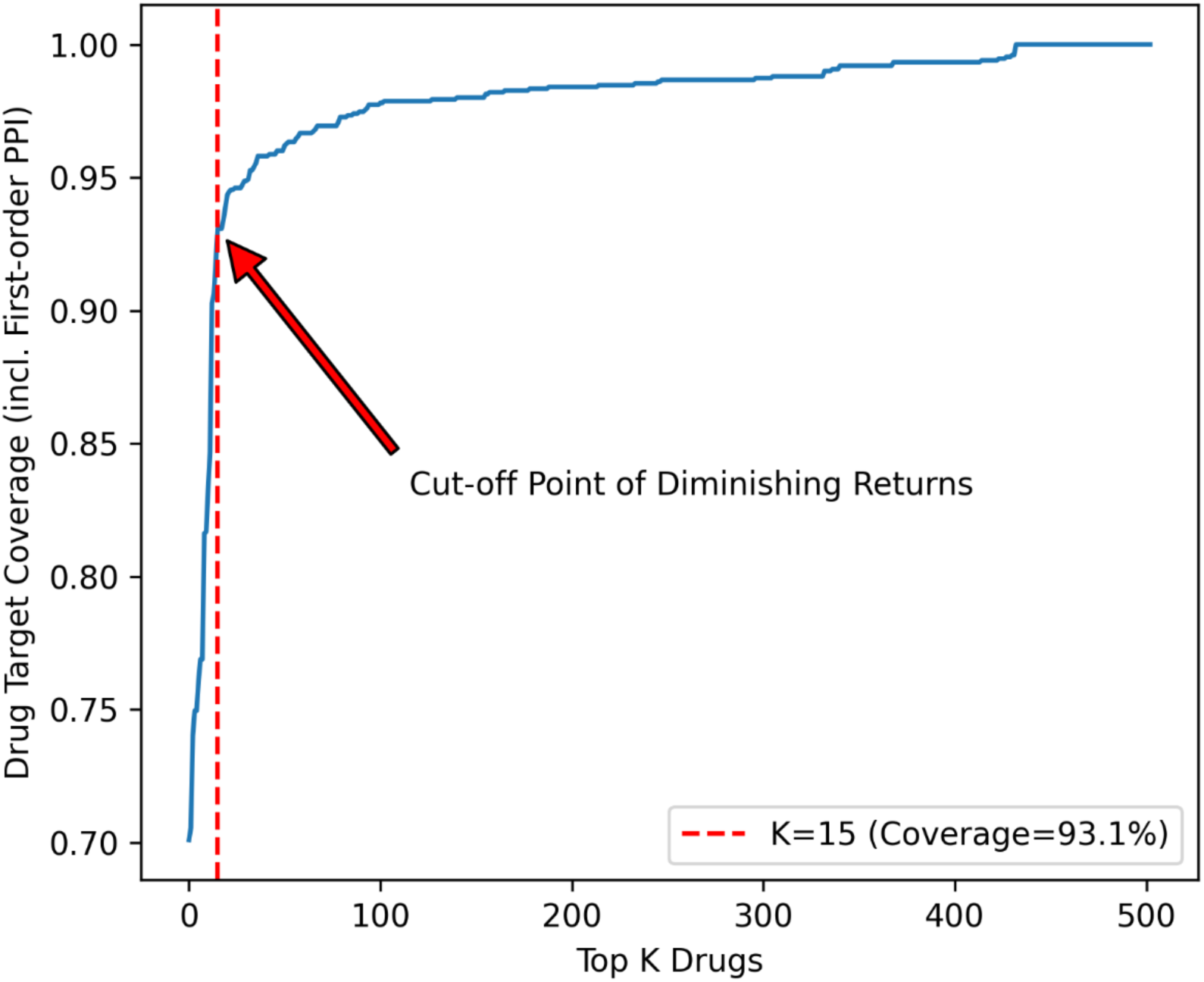
The Cut-off Point to Determine the Number of Top Drug Candidates

Based on the top 15 drug candidates, we selected a lead combination more probable to hit AD-associated genes than others and achieve the maximum synergistic effect. Specifically, we calculated the DeepDrug score of drug combinations (from 2 to *n* drugs). The optimal number of drugs in drug combinations was determined by the point when the lead drug combination received the maximum DeepDrug score. The DeepDrug score of a drug combination was calculated by merging individual drug nodes into super-drug nodes in the biomedical graph, accounting for (i) the DeepDrug scores of individual drug candidates and their pairwise interactions, (ii) the chemical structures of individual drug candidates and their pairwise interactions, and (iii) the shared and complementary drug targets. The super-drug nodes, super-drug target edges, and super-drug-drug edges were constructed and weighted as follows.

*Super-Drug Nodes:* Each super-drug node represented one drug combination consisting of multiple drugs. Specifically, if the DeepDrug score can measure a single drug’s AD treatment effect (ranging from 0 to 1), the weight of an *n*-drug combination, indicating the expected combination effect, was calculated as follows. First, the weight of a two-drug combination (i.e., the pairwise effect) was calculated based on the Bliss model, a commonly used reference model for two-drug combinations^98^:

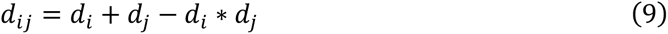

where

**d*_ij_* represents the weight of one drug pair {*i*, *j*}, and *d_i_* represents the DeepDrug score of one drug *i*.

Second, for a drug combination consisting of more than two drugs, the weight was calculated based on a mechanism-independent reference model, utilizing an Isserlis-like formula considering pairwise effects only while ignoring high-order interactions^116^. The original formulas proposed in Wood et al. (2012)^116^ focused on three-drug and four-drug combinations and were extended to *n*-drug combinations here:

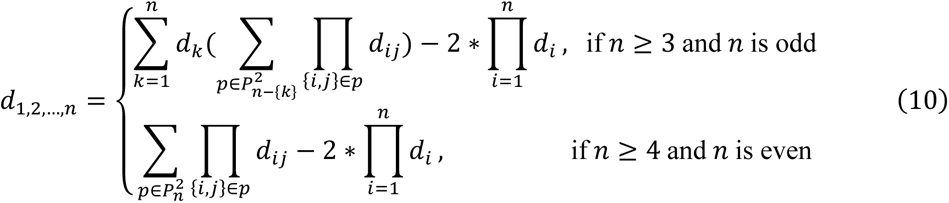

where

*d*_1,2,…,n_ represents the weight of an *n*-drug combination,

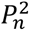 represents all unique ways of partitioning {1,2, … , *n*} into pairs,

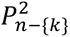 represents all unique ways of partitioning {1,2, … , *n*} without the element *k* into pairs,

*d_ij_* represents the weight of one drug pair {*i*, *j*}, and *d_i_* represents the DeepDrug score of one drug *i*.

For example, the weight of a three-drug combination was: *d*_1,2,3_ = *d*_1_ ∗ *d*_23_ + *d*_2_ ∗ *d*_13_ + *d*_3_ ∗ *d*_12_ − 2 ∗ *d*_1_ ∗ *d*_2_ ∗ *d*_3_), and the weight of a four-drug combination was: *d*_1,2,3,4_ = *d*_12_ ∗ *d*_34_ + *d*_13_ ∗ *d*_24_ + *d*_14_ ∗ *d*_23_ − 2 ∗ *d*_1_ ∗ *d*_2_ ∗ *d*_3_ ∗ *d*_4_.

In addition, the super-drug node’s high-dimensional weight vector was calculated based on each drug node’s high-dimensional weight vector. The intuition was that one part of a drug’s chemical structure could complement another drug and the molecular representation of a drug combination could be related to its combination effect. Each drug’s weight vector was represented by its fingerprint (bit string), capturing different fragments of the drug’s molecular structure, and enabling zero-shot prediction for drug combination activity with previously unseen molecular information^117^. Specifically, for a two-drug combination, the same Bliss-based formula^98^ was applied to single-drug fingerprints to calculate the corresponding combination’s fingerprint: *fp_ij_* = *fp_i_* + *fp_j_* – *fp_i_* ⊗ *fp_j_*, where ⊗ denotes element-wise multiplication. Similarly, for an *n*-drug combination with more than two drugs, the same Isserlis-like formula^116^ was applied to single-drug fingerprints to calculate the corresponding combination’s fingerprint. For example, for a three-drug combination, its fingerprint was calculated as follows: *fp*_1,2,3_ = *fp*_1_ ⊗ *fp*_23_ + *fp*_2_ ⊗ *fp*_13_ + *fp*_3_ ⊗ *fp*_12_ − 2 ∗ *fp*_1_ ⊗ *fp*_2_ ⊗ *fp*_3_), where ⊗ denotes element-wise multiplication. Finally, the SVD method was used to compress each super-drug fingerprint into a fixed-length vector.

*Super-Drug Target Edges:* Each super-drug-target edge was constructed based on the interactions between the corresponding single drugs and their targets. As a result, a super-drug node was more likely to cover more diverse targets. The weight of super-drug-target edges were calculated by summing the corresponding single drug-target edge weights (normalized to [1, 2]) while considering their signs. The sign of super-drug-target edges were determined based on the sign of aggregated weights. The rationale was to account for the additive effects (e.g., an increase in the total effect if two drugs inhibited the same protein) while addressing the contradicted effects (e.g., a decrease in the total effect if one drug activated a target while another inhibited the same target).

*Super-Drug-Drug Edges:* All super-drug-drug edges (i.e., interactions between the super-drug nodes) were ignored, given that higher-order drug-drug interactions could often be ignored when modeling their combination effects^116^ and addressing the side effects of taking a combination of drug combinations were beyond the scope of identifying a lead drug combination.

Finally, we selected a super-drug node (i.e., a lead combination of drugs) achieving the best score. We evaluated the drug combinations selected from the top 15 drugs, ranging from two-drug combinations to *n*-drug combinations. The detailed procedure is shown in Algorithm 2.

#### Algorithm 2. Lead Drug Combination Selection

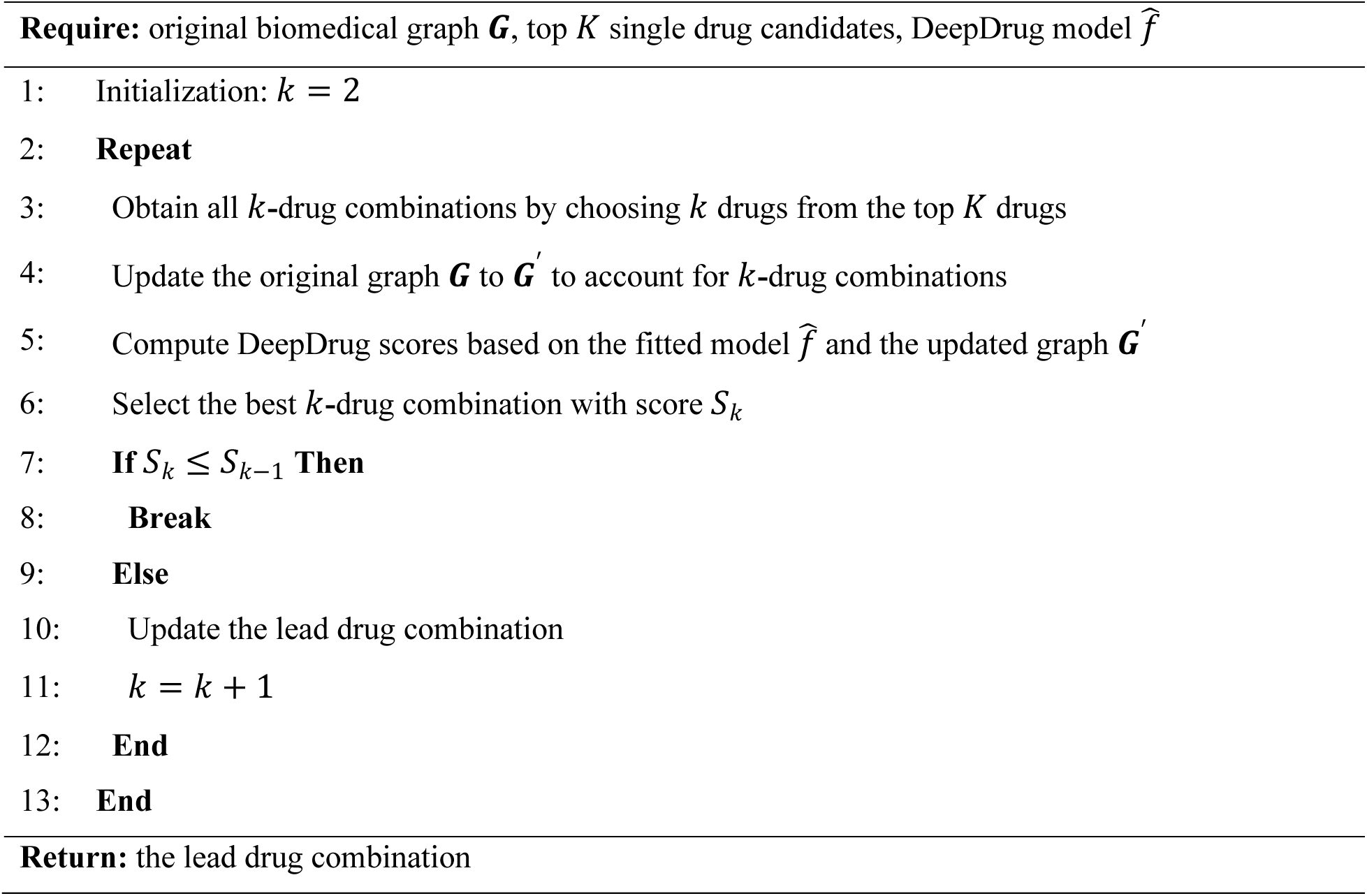

## Data Availability

The drug and drug-drug interaction data were obtained from DrugBank (https://go.drugbank.com). The drug target data were obtained from DrugBank (https://go.drugbank.com), DrugCentral (https://drugcentral.org), ChEMBL (https://www.ebi.ac.uk/chembl/), and BindingDB (https://www.bindingdb.org/). The protein-protein interaction and pathway data were obtained from STRING (https://string-db.org).

## Additional Information

This work including contribution of six of the co-authors is under patent protection (University of Hong Kong and Tel Aviv University).

## Acknowledgments

This research was supported in part by the US National Academy of Medicine (NAM) Healthy Longevity Catalyst Award 2021 and “An Artificial Intelligence (AI)-driven Causal Approach for Early Diagnosis and Treatment of Late Onset Alzheimer’s Disease” under Seed Fund for Collaborative Research, The University of Hong Kong, June 2023. The authors thank Dr. Ruiqiao Bai of the University of Hong Kong for assisting with part of the literature review.

## Data Availability

The drug and drug-drug interaction data were obtained from DrugBank (https://go.drugbank.com). The drug target data were obtained from DrugBank (https://go.drugbank.com), DrugCentral (https://drugcentral.org), ChEMBL (https://www.ebi.ac.uk/chembl/), and BindingDB (https://www.bindingdb.org/). The protein-protein interaction and pathway data were obtained from STRING (https://string-db.org). The preprocessed biomedical network datasets, including genes and their weights (Table S1), drugs and their weights (Table S2), proteins/targets and their weights (Table S3), and edges and their weights (Tables S4-7), are available in Supplementary Data.

## Author Contributions

VL and JL developed the overall framework and identified the key novelties and significance and are the main drivers of this work. They also obtained research funding. YH, QZ, VL, JL designed, and YH, QZ and TK operationalized the GNN model and the biomedical graph, TK and YH collected data on nodes and edges of the biomedical graph, YH, TK, VL and JL wrote the paper, JL, VL and YH revised all drafts and finalized the manuscript. YH prepared all figures. IG and JD provided the domain-specific expert knowledge on the biomedical graph and IG initiated the idea of drug combinations. VL and JL are the senior and the corresponding authors of the funded project and provide the funding support for this study.

